# Genomic Epidemiology of Multi-Modal ESBL Gene Transmission among Enterobacterales in a Neonatal Unit

**DOI:** 10.64898/2026.07.03.26357218

**Authors:** Melody J Parker, Katie Hopkins, Kevin Chau, Jack Cregan, Sarah Oakley, Lucinda Barrett, Katie Jeffrey, Lisa Butler, Stephane Paulus, Bernadette Young, David W Eyre, Philip W Fowler, Nicole Stoesser, Nicholas Sanderson, Philip Bejon

## Abstract

Antimicrobial resistance genes (ARGs) can spread via horizontal transfer or clonal expansion. We investigated the genomic epidemiology of extended-spectrum beta-lactamase (ESBL)–producing *Klebsiella pneumoniae* and *Escherichia coli* in a neonatal unit. Between January and November 2023, 53 ESBL isolates were obtained from 23 neonates via routine screening and clinical sampling. Long-read nanopore sequencing identified *bla*_CTX-M-15_ as the dominant ESBL gene, alongside *bla*_CTX-M-65_, and *bla*_CTX-M-27_. Among 49 *bla*_CTX-M-15_ isolates, 33 carried the gene on plasmids and in the remainder it was located on the chromosome. ESBL isolates belonged to one of seven MLST sequence types; *E. coli* isolates were dominated by ST131/ST131-like/ST5640 lineages, while *K. pneumoniae* were predominantly ST13. Plasmids (ESBL and non-ESBL-associated) clustered into 18 communities, five of which contained plasmid bearing *bla*_CTX-M_ genes. The largest cluster comprised IncFIB(K) plasmids from *K. pneumoniae* ST13, although, these were predicted as “non-mobilizable” by MOB-suite, and belonged to isolates from a clonally disseminated strain. No evidence of *bla*_CTX-M_ dissemination via shared plasmids was identified. Meanwhile, chromosomal phylogenetic analysis identified four distinct clonal clusters with ≤7 SNP differences (n=10, 5, 5, and 2 patients). In this setting, genomic analysis supported clonal dissemination of several *bla*_CTX-M_-associated strains as the main outbreak mechanism, affecting 20/23 neonates, rather than plasmid-mediated transmission.

## Introduction

Antimicrobial resistance (AMR) is a major global health threat, with hospitals being key AMR hotspots [1–3]. Healthcare settings can create highly conducive conditions for the spread of bacteria due to significant selection pressures such as high rates of antimicrobial use, the presence of vulnerable patients, interaction with healthcare professionals who can also carry these organisms, and environmental contamination [4, 5]. Neonates are especially susceptible to bacterial infections, due to their immature immune systems [6, 7]. In the United Kingdom, approximately one in seven newborn infants are admitted to neonatal intensive care units (NICUs). Within these settings, the frequent use of invasive indwelling devices further contributes to the risk of severe nosocomial infections [8]. Healthcare-associated infections in neonates are associated with higher mortality rates, extended hospital stays, increased healthcare costs, and a greater likelihood of neurodevelopmental disabilities among survivors [9]. While antibiotics are essential for the management of neonatal infections, the increasing prevalence of antimicrobial resistance threatens their efficacy, a particularly critical concern in NICUs where therapeutic alternatives are limited and the patients are highly vulnerable.

AMR can arise via a diverse range of genetic mechanisms, all of which limit the efficacy of treatment [10]. The mechanisms by which antimicrobial-resistance genes (ARGs) confer resistance and spread through bacterial populations are frequently complex, leading to different patterns of evolution and transmission in outbreaks. In some instances, the genetic basis of resistance is straightforward; for example, when a small number of mutation- or gene-based changes contribute clearly to an AMR phenotype, and are disseminated vertically in a clonal fashion leading to a strain-based outbreak. In other instances, such as those seen in third-generation cephalosporin resistance in Enterobacterales, resistance is more complicated, as it can be encoded by multiple mechanisms and frequently disseminated both vertically and horizontally via mobile genetic elements (MGEs), including plasmids and transposons [11]. This nested “Russian-doll”-like resistance hierarchy can significantly complicate both surveillance and Infection Prevention and Control (IPC) efforts, and has led to the need for new strategies to evaluate gene-based outbreaks [12].

Plasmid-mediated transmission is a key mechanism underlying the spread of AMR in complex outbreaks. Identification of incompatibility replicon types, relaxase genes, AMR genes and mobility genes have long been used to help classify plasmids. For investigating putative plasmid-driven outbreaks, detailed characterisation of plasmid structure is essential, as the presence of shared target genes alone cannot distinguish recent plasmid transmission from common ancestry. Unlike clonal outbreak investigations, which often rely on single nucleotide polymorphism (SNP)-based analysis, plasmid outbreak investigations must also consider structural recombination due to the high recombination rate of plasmids. It is only more recently with the development of long-read sequencing that we have been able to effectively resolve whole plasmid genomes, with the longer reads spanning over repeat regions that commonly appear on plasmids, e.g. insertion sequences and transposons. Due to the lower accuracy of Oxford Nanopore Technologies plc (ONT) sequencing and the high costs of Pacific Biosciences, Inc. sequencing, this method of obtaining the plasmid stucture has often been complimented by more accurate (and cheaper) short reads (e.g. Illumina, Inc.) to improve the overall accuracy of the detected SNPs. However, ONT accuracy has improved substantially in recent years [13], with a recent study reporting that Enterobacterales assemblies can be equivalent in quality to hybrid/Illumina assemblies [14].

To understand plasmid relatedness, plasmid clustering tools such as Pling have been developed; Pling estimates pairwise containment distances via an alignment-based method which groups plasmids into communities, calculating the number of double-cut and joins (DCJ) between samples to cluster the plasmids in these communities into subcommunities, thereby estimating how many recombination events have happened as a proxy for relatedness. However, one should not assume that every plasmid cluster reflects recent transmission; these may instead represent less recent transfer events since some plasmids, for example IncL plasmids [15], can remain stable over months to years. In addition, plasmids associated with a clonal outbreak are likely to cluster together.

Recently, several studies have used long-read sequencing to demonstrate that shared, mobilisable plasmids can disseminate antimicrobial resistance genes across strains and species; this phenomenon is well documented in carbapenemase-producing Enterobacterales (CPEs), where plasmids are recognised as key drivers of multispecies outbreaks in healthcare settings[16–20]. For ESBL-producing Enterobacterales, however, evidence for plasmid-mediated dissemination remains limited. Notably, Pedersen *et al.* [21] identified a *bla*_CTX-M-15_-positive IncFIIK5/IncR plasmid disseminated across 48 sequence types of *K. pneumoniae* in both hospitalised and community-associated isolates in Tanzania. Consistent with this, both *in vitro* and *in vivo* studies provide evidence for horizontal transfer of ESBL genes across strains and species [22–24], with antibiotics shown to induce conjugation of ESBL-bearing plasmids [25]. Together, these studies show that whole ESBL-gene-bearing plasmid transfer between strains and species is biologically plausible, but possibly under-characterised in the literature. Given the documented occurrence of plasmid-mediated CPE outbreaks and the fact that our ESBL-producing isolates are from two different species, we investigated whether ESBL gene transmission was driven by a shared plasmid in our case.

In an NICU in Oxfordshire, UK in 2023, multiple isolates from neonates colonised or infected with ESBL-positive E. coli and K. pneumoniae were flagged through IPC surveillance based on epidemiological outbreak definitions (i.e. an increase over the baseline number of positives). The detection of ESBL-producing isolates from multiple species prompted an investigation into whether plasmid-mediated horizontal gene transfer had occurred within the NICU. We therefore explored whether a similar plasmid-mediated mechanism was responsible for the increase in ESBL-producing isolates in this setting. To address this, we applied ONT long-read sequencing to fully characterise the genomic and plasmid context of these isolates. This is the first study, in our knowledge, to use ONT as the sole genetic sequencing method to investigate the spread of ESBL-producing Enterobacterales in a neonatal ward. Particular emphasis was placed on resolving the structure of plasmids and their potential contribution to the dissemination of resistance genes, leading to a deeper understanding of the mechanisms underlying spread. Together, our approach provides detailed insight into the transmission dynamics that occurred within the Oxfordshire NICU and could inform future IPC decision-making.

## Results

### ESBL-producing *E. coli* or *K. pneumoniae* were identified in 8.5% of screened neonates in 2023

A total of 7,980 isolates were collected from 837 neonatal patients between 01.01.2023 to 31.12.2023 (Figure 1. A minority (303, 3.8 %) of these isolates obtained from 123 patients were classified as containing Enterobacterales, with 164 *E. coli* from 89 patients, 78 *K. pneumoniae* from 18 patients, and 61 isolates from 34 patients were classified as other or non-specific species. Within this subset, a majority of *E. coli* (112/164, 68.3 %) and *K. pneumoniae* (69/78, 88.5 %) isolates were identified as ESBL-producers. These ESBL-producing *E. coli* and *K. pneumoniae* isolates were identified via two routes: most isolates (149/181, 82.3%) were acquired from an ESBL rectal screening programme which collected 1,634 isolates from 656 patients; the remainder (32/181, 17.7 %) were obtained from routine diagnostic testing. Overall, 56 of 656 patients screened (8.5 %) had at least one positive result for ESBL-producing *E. coli* or *K. pneumoniae*. The remaining 32 ESBL-producing *E. coli* or *K. pneumoniae* isolates, from 13 patients, originated from cultures derived from surface swab (n=11), blood (n=9), respiratory (n=8), urine (n=2), carbapenemase (CPE) screen (n=1), and pus (n=1) clinical diagnostic specimens.

**Figure 1:**
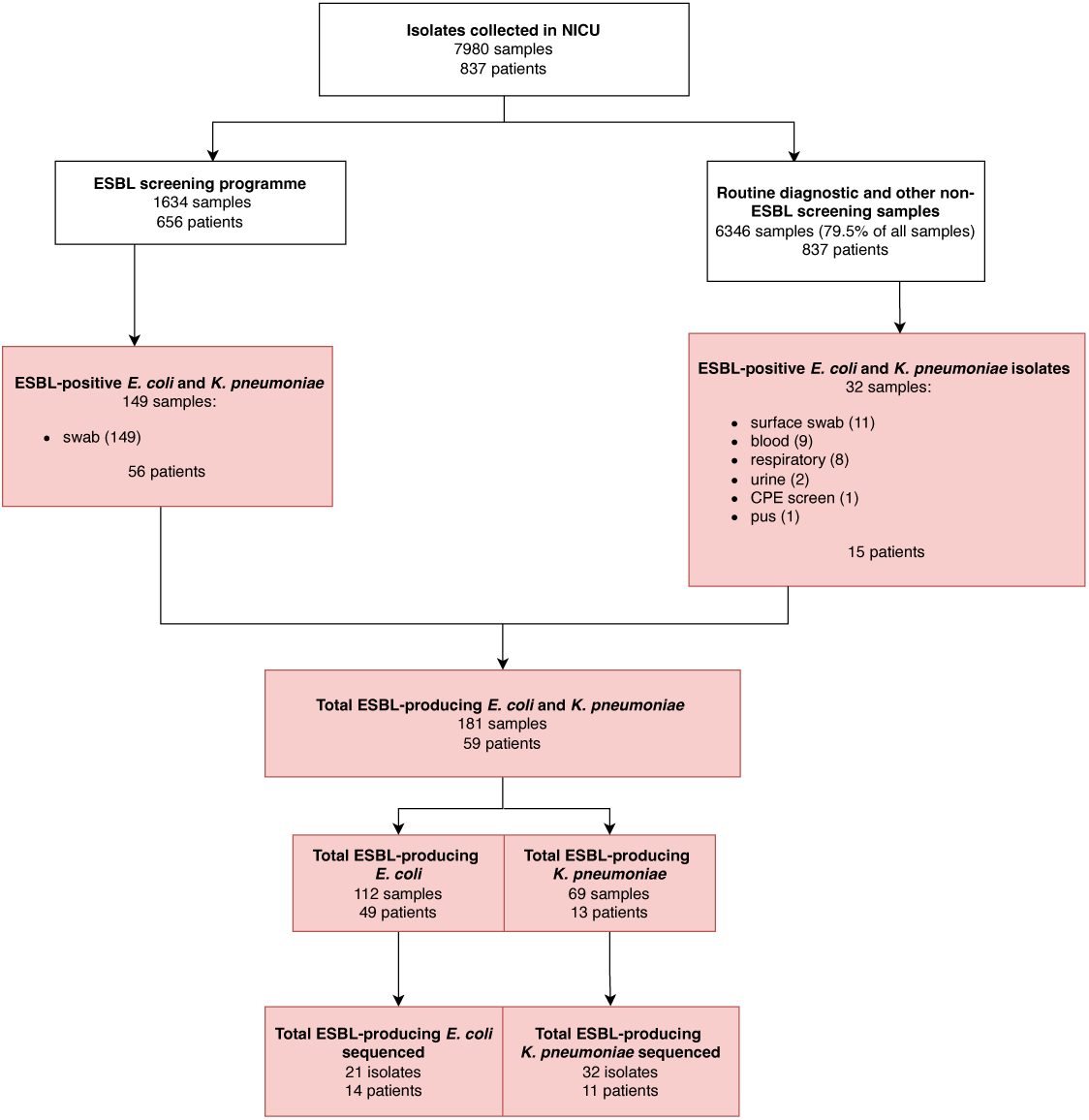
Isolate collection routes of ESBL-producing *K. pneumoniae* and *E. coli*. Flowchart showing the number of ESBL-positive *E. coli* and *K. pneumoniae* isolates collected from patients in Oxfordshire NICU in 2023, and their sampling routes. Records were extracted from the IORD database.

### ESBL-positive isolates displayed substantial genomic diversity

In total, 53 ESBL-producing neonatal isolates were sequenced (out of the 181 ESBL-positive *E. coli* and *K. pneumoniae* isolates identified in the study period [29.3%]), comprising 32 *K. pneumoniae* and 21 *E. coli* isolates based on phenotypic identification. Isolates were collected during two distinct periods: 42 isolates between 02.01.23 and 23.04.23, and 11 isolates between 07.10.23 and 20.11.23 (Figure S1).

A *bla*_CTX-M_ ESBL antimicrobial resistance gene (ARG) was identified in all sequenced isolates (Figure 2). No non-*bla*_CTX-M_ ESBL genes were identified. Three *bla*_CTX-M_ variants were identified by AMRfinder (all with 100% coverage of, and identical to, the reference allele): these were *bla*_CTX-M-15_ (n=49 isolates), *bla*_CTX-M-65_ (n=2), or *bla*_CTX-M-27_ (n=2) (Table 1). A majority (37/53, 70 %) of isolates had the *bla*_CTX-M_ allele located on plasmids whilst for the remaining 16 the gene was integrated in the chromosome. Isolates belonged to one of seven different multilocus sequence type (MLST) groups; those containing *E. coli* belonged to ST131/ST131-like/ST5640 (n=12/21, 57.1%), ST127 (n=5) or ST1193 (n=4), while most *K. pneumoniae* isolates belonged to ST13 (n=30/32, 93.7 %), with single isolates representing ST1589 and ST307. ST131-like isolates were defined as sharing six of seven MLST alleles with ST131, with the remaining allele being a novel full-length variant closely related to the corresponding ST131 allele at that locus. ST5640, while classified as a distinct MLST sequence type, also shares six of seven alleles with ST131, with the final allele differing by only one SNP (Table S2). The presence of multiple MLST backgrounds in both *E. coli* and *K. pneumoniae* indicated that the isolates did not all belong to a single clonal outbreak for each species. The presence of three distinct *bla*_CTX-M_ variants indicated at least three independent introductions of *bla*_CTX-M_ alleles into the ward.

**Figure 2:**
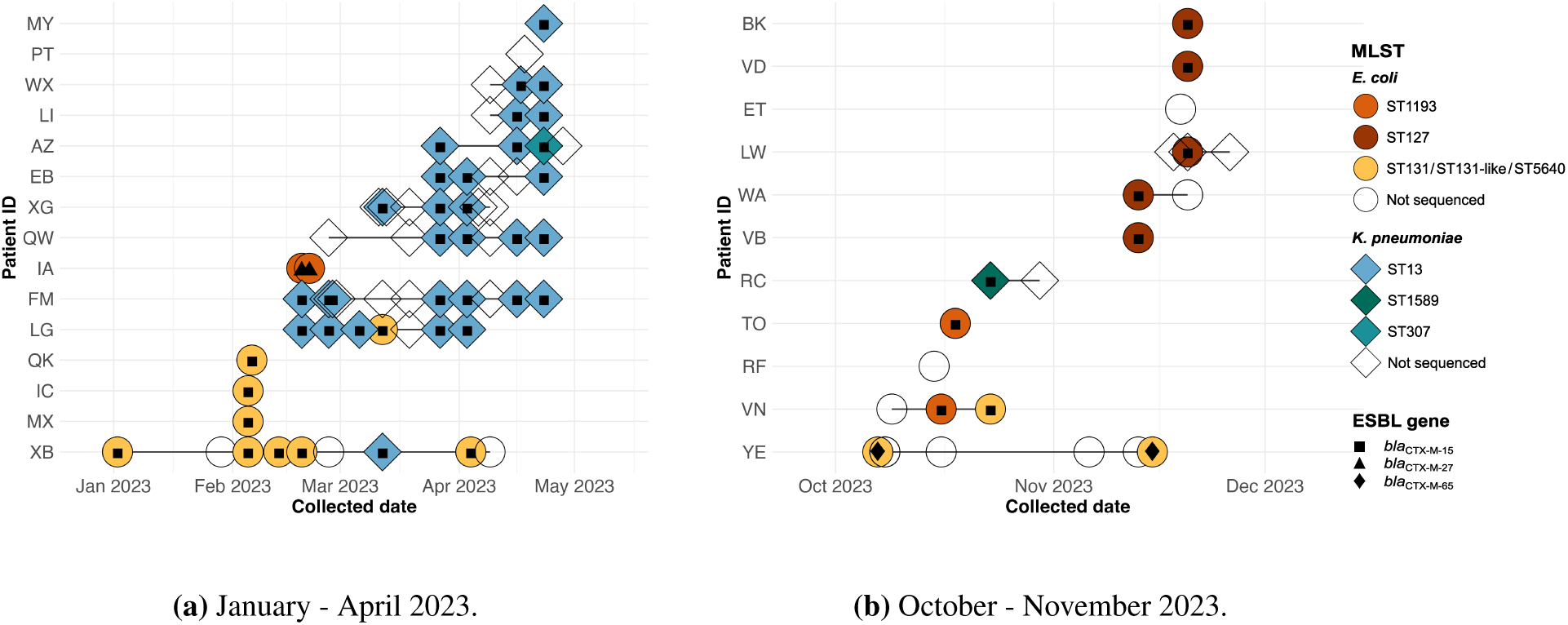
Time-series plot of neonatal isolates collected in 2023 and flagged by IPC. For the isolates that were sequenced, *E. coli* are represented by circular and warm-coloured points; *K. pneumoniae* are represented by diamond-shaped and cool-coloured points. The points of unsequenced isolates are shown using the same species-specific point shape, but are transparent. (a) The first batch, from January - April 2023. (b) The second batch, from October - November 2023. Patient initials were randomised for confidentiality.

**Table 1:** Per-patient *E. coli* and *K. pneumoniae* MLST sequence types and *bla*_CTX-M_ variants.

| Patient ID | Total samples | *E. coli* |  | *K. pneumoniae* |  |
| --- | --- | --- | --- | --- | --- |
|  |  | MLST (n) | *bla*<sub>CTX-M</sub> variant | MLST (n) | *bla*<sub>CTX-M</sub> variant |
| XB | 6 | ST131-like<br>ST131 (4) | *bla*<sub>CTX-M-15</sub><sup>*</sup> (5) | ST13 | *bla*<sub>CTX-M-15</sub><sup>†</sup> |
| MX | 1 | ST131-like | *bla*<sub>CTX-M-15</sub><sup>*</sup> | — | — |
| IC | 1 | ST5640 | *bla*<sub>CTX-M-15</sub><sup>*</sup> | — | — |
| QK | 1 | ST131 | *bla*<sub>CTX-M-15</sub><sup>*</sup> | — | — |
| LG | 6 | ST131 | *bla*<sub>CTX-M-15</sub><sup>*</sup> | ST13 (5) | *bla*<sub>CTX-M-15</sub><sup>†</sup> (5) |
| FM | 7 | — | — | ST13 (7) | *bla*<sub>CTX-M-15</sub><sup>†</sup> (7) |
| IA | 2 | ST1193 (2) | *bla*<sub>CTX-M-27</sub><sup>†</sup> (2) | — | — |
| QW | 4 | — | — | ST13 (4) | *bla*<sub>CTX-M-15</sub><sup>†</sup> (4) |
| XG | 3 | — | — | ST13 (3) | *bla*<sub>CTX-M-15</sub><sup>†</sup> (3) |
| EB | 3 | — | — | ST13 (3) | *bla*<sub>CTX-M-15</sub><sup>†</sup> (3) |
| AZ | 3 | — | — | ST13 (2)<br>ST307 | *bla*<sub>CTX-M-15</sub><sup>†</sup> (3) |
| LI | 2 | — | — | ST13 (2) | *bla*<sub>CTX-M-15</sub><sup>†</sup> (2) |
| WX | 2 | — | — | ST13 (2) | *bla*<sub>CTX-M-15</sub><sup>†</sup> (2) |
| MY | 1 | — | — | ST13 | *bla*<sub>CTX-M-15</sub><sup>†</sup> |
| YE | 2 | ST131 (2) | *bla*<sub>CTX-M-65</sub><sup>†</sup> (2) | — | — |
| VD | 1 | ST127 | *bla*<sub>CTX-M-15</sub><sup>*</sup> | — | — |
| VB | 1 | ST127 | *bla*<sub>CTX-M-15</sub><sup>*</sup> | — | — |
| BK | 1 | ST127 | *bla*<sub>CTX-M-15</sub><sup>*</sup> | — | — |
| VN | 2 | ST1193<br>ST131 | *bla*<sub>CTX-M-15</sub><sup>*</sup><br>*bla*<sub>CTX-M-15</sub><sup>†</sup> | — | — |
| LW | 1 | ST127 | *bla*<sub>CTX-M-15</sub><sup>*</sup> | — | — |
| RC | 1 | — | — | ST1589 | *bla*<sub>CTX-M-15</sub><sup>†</sup> |
| WA | 1 | ST127 | *bla*<sub>CTX-M-15</sub><sup>*</sup> | — | — |
| TO | 1 | ST1193 | *bla*<sub>CTX-M-15</sub><sup>*</sup> | — | — |

Patient-level summary of sequenced *E. coli* and *K. pneumoniae* isolates showing MLST sequence types and associated *bla*<sub>CTX-M</sub> variants. Patient initials (Patient ID) are anonymised. Since some patients were sampled multiple times, the number in parentheses details the number of the samples containing the MLST sequence type or *bla*<sub>CTX-M</sub> allele. \* *bla*<sub>CTX-M</sub> gene located on the chromosome. † *bla*<sub>CTX-M</sub> gene located on a plasmid.

All *K. pneumoniae* isolates carried their *bla*_CTX-M_ gene on plasmids, whereas the *E. coli* isolates showed a mixture of plasmid- and chromosomal localisation. *bla*_CTX-M-15_ was carried on both chromosomes and plasmids, whereas *bla*_CTX-M-27_ and *bla*_CTX-M-65_ were exclusively plasmid-borne.

Among isolates in which *bla*_CTX-M_ was plasmid-borne, both *bla*_CTX-M-15_ and *bla*_CTX-M-27_ were identified in multiple patients, while *bla*_CTX-M-65_ was detected in two *E. coli* ST131 isolates in a single patient, with no indication of within-patient transfer. Together, these observations motivated subsequent analyses to assess whether dissemination of *bla*_CTX-M-15_ and *bla*_CTX-M-27_ occurred via variant-specific shared plasmids across isolates.

### *bla*_CTX-M_ variants occur in diverse genomic contexts

All *K. pneumoniae* isolates carried an IncFIB(K)-type replicon and all *K. pneumoniae* ST13 isolates carried *bla*_CTX-M-15_ on a plasmid with an IncFIB(K)(pCAV1099-114) replicon. In the other two *K. pneumoniae* isolates, the ST1589 isolate carried *bla*_CTX-M-15_ on a plasmid containing both IncFIB(K) and IncFII(K) replicons, whereas the ST307 isolate carried *bla*_CTX-M-15_ on a plasmid with an IncFIB(K) replicon only; notably, these two IncFIB(K) replicons are distinct from the IncFIB(K)(pCAV1099-114) replicon identified in ST13 plasmids. The only *E. coli* plasmid (151,122 bp) carrying a *bla*_CTX-M-15_ gene belonged to an ST131 isolate, and contained replicons IncFIA, IncFIB(AP001918) and IncFII(pRSB107). In *E. coli*, all other *bla*_CTX-M-15_ were carried on the chromosome: six ST131, two ST131-like, one ST5640, two ST1193, and all five ST127 isolates. Both *bla*_CTX-M-27_–positive isolates were *E. coli* ST1193. Two isolates collected in February 2023 from patient IA carried *bla*_CTX-M-27_ on a plasmid containing the Col156, IncFIA and IncFIB(AP001918) replicons. Both instances of *bla*_CTX-M-65_ were associated with a plasmid with replicon types IncHI2 and IncHI2A and were taken from a single patient (YE). All 49 examples of *bla*_CTX-M-15_ were found on either a chromosome or an IncFIB plasmid.

### *bla*_CTX-M_-bearing-plasmids clustered into five communities

Given the diversity of host species and MLST sequence types observed, together with the predominance of the *bla*_CTX-M-15_ allele, plasmid clustering was performed to assess whether ESBL transmission could be attributed to a common shared plasmid. Pling was used to group all plasmids that assembled into complete circular sequences, resulting in 18 plasmid communities (labelled C1–C18) based on their containment distance (Table 2 and Figures S8, S9, S10, S11, S12, S13). Node compression was performed to simplify plots, further clustering plasmids where DCJ-distance was zero. Five of the 17 plasmid communities (C2, C7, C15, C17 & C18) contained at least one plasmid bearing a *bla*_CTX-M_ gene.

**Table 2:** Plasmid communities.

| Community ID | No. of plasmids | No. of host species | No. of MLSTs | No. of patients | *bla*<sub>CTX-M</sub> variants | Sampling months (2023) |
| --- | --- | --- | --- | --- | --- | --- |
| community 1 | 25 | 2 | 6 | 16 | - | APR, FEB, JAN, MAR, NOV, OCT |
| community 2 | 28 | 1 | 1 | 10 | *bla*<sub>CTX-M-15</sub> (28) | APR, FEB, MAR |
| community 3 | 31 | 2 | 5 | 14 | - | APR, FEB, MAR |
| community 4 | 25 | 2 | 5 | 13 | - | APR, FEB, JAN, MAR |
| community 5 | 2 | 1 | 1 | 1 | - | NOV, OCT |
| community 6 | 14 | 2 | 5 | 9 | - | APR, FEB, JAN, MAR |
| community 7 | 18 | 1 | 5 | 12 | *bla*<sub>CTX-M-27</sub> (2)<br>*bla*<sub>CTX-M-15</sub> | APR, FEB, JAN, MAR, NOV, OCT |
| community 8 | 2 | 1 | 1 | 1 | - | NOV, OCT |
| community 9 | 5 | 2 | 4 | 4 | - | FEB, JAN, MAR, OCT |
| community 10 | 3 | 2 | 2 | 2 | - | FEB |
| community 11 | 2 | 1 | 1 | 1 | - | NOV, OCT |
| community 12 | 2 | 1 | 1 | 1 | - | NOV, OCT |
| community 13 | 2 | 1 | 1 | 2 | - | OCT |
| community 14 | 4 | 1 | 1 | 3 | - | FEB, OCT |
| community 15 | 2 | 1 | 1 | 1 | *bla*<sub>CTX-M-65</sub> (2) | NOV, OCT |
| community 16 | 1 | 1 | 1 | 1 | - | OCT |
| community 17 | 1 | 1 | 1 | 1 | *bla*<sub>CTX-M-15</sub> | APR |
| community 18 | 1 | 1 | 1 | 1 | *bla*<sub>CTX-M-15</sub> | OCT |

Since some plasmid communities contained multiple *bla*<sub>CTX-M</sub>-bearing plasmids, the number in parentheses details the number of the plasmids containing the *bla*<sub>CTX-M</sub> allele.

**Table 3:**
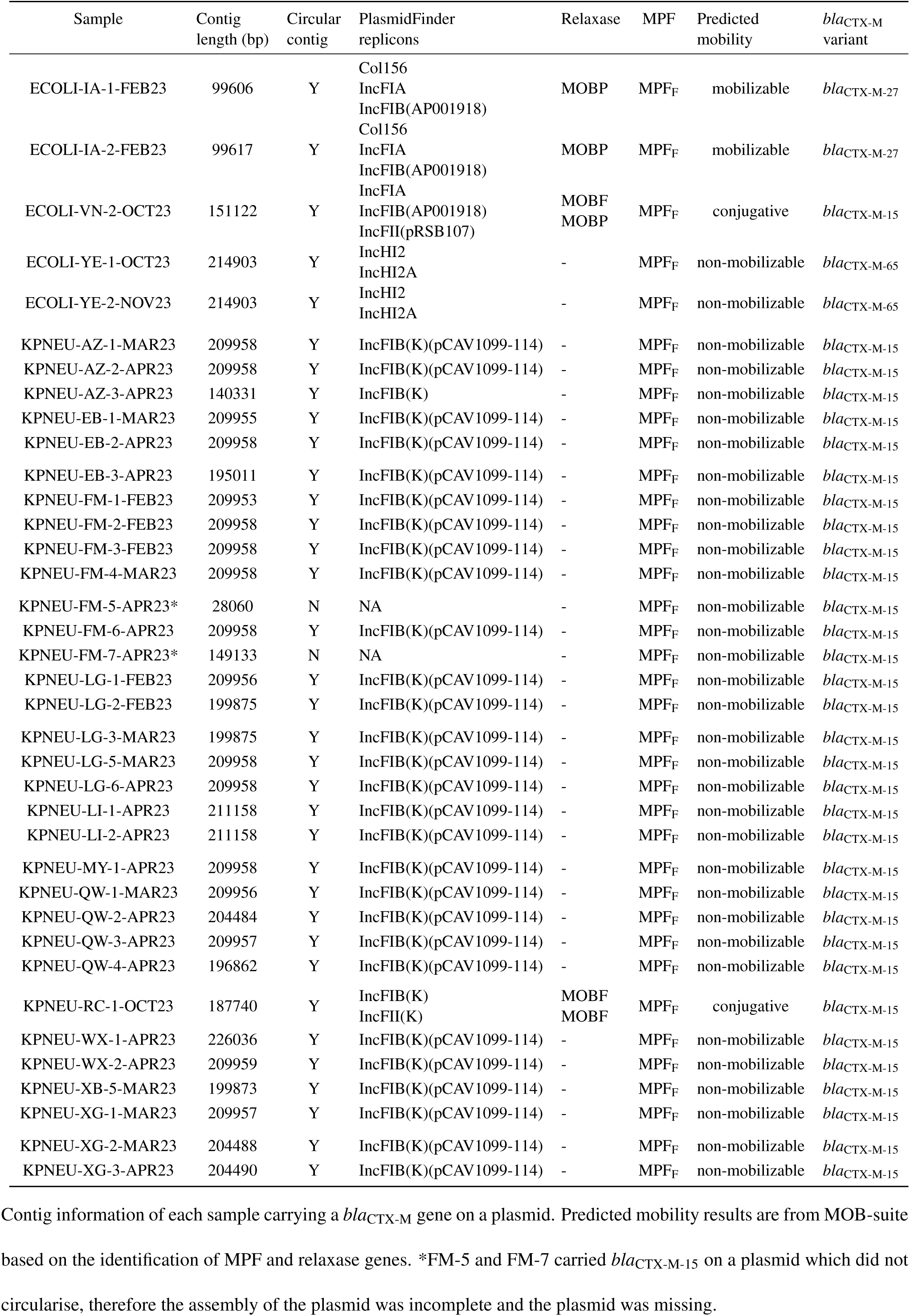
Plasmid-associated contigs carrying ESBL genes.

The largest of these five communities, C2, consisted exclusively of plasmids that carried an IncFIB(K)(pCAV1099-114) replicon all of which were carried by *K. pneumoniae* ST13 isolates and encoded *bla*_CTX-M-15_ (Figure 3b). All of these plasmids were predicted by MOB-suite to be non-mobilisable (Figure S8b), due to the absence of identifiable relaxase or mating-pair formation (MPF) genes, and exhibited a maximum pairwise DCJ-distance of three, indicating a high degree of structural similarity. This community contained all circularised *K. pneumoniae* ST13 plasmids carrying *bla*_CTX-M-15_ included in the analysis (30/32), and no plasmids from any other MLST. The remaining two ST13 *bla*_CTX-M_-containing contigs were not included in the plasmid clustering analysis due to their fragmented assembly that precluded plasmid circularisation; however, in both cases one contig from the assemblies of these plasmids carried a *bla*_CTX-M-15_ gene and another contig carried an IncFIB(K)(pCAV1099-114) replicon, suggesting that these likely represent the same plasmid type as those in C2 (Figure S7). Together, these features indicate the outbreak contains a highly conserved, non-mobilisable *bla*_CTX-M-15_-harbouring plasmid associated with *K. pneumoniae* ST13 isolates. Given the predicted lack of mobilisation and the restriction of this plasmid to a single MLST sequence type, further investigation of host genomic relatedness was warranted.

**Figure 3:**
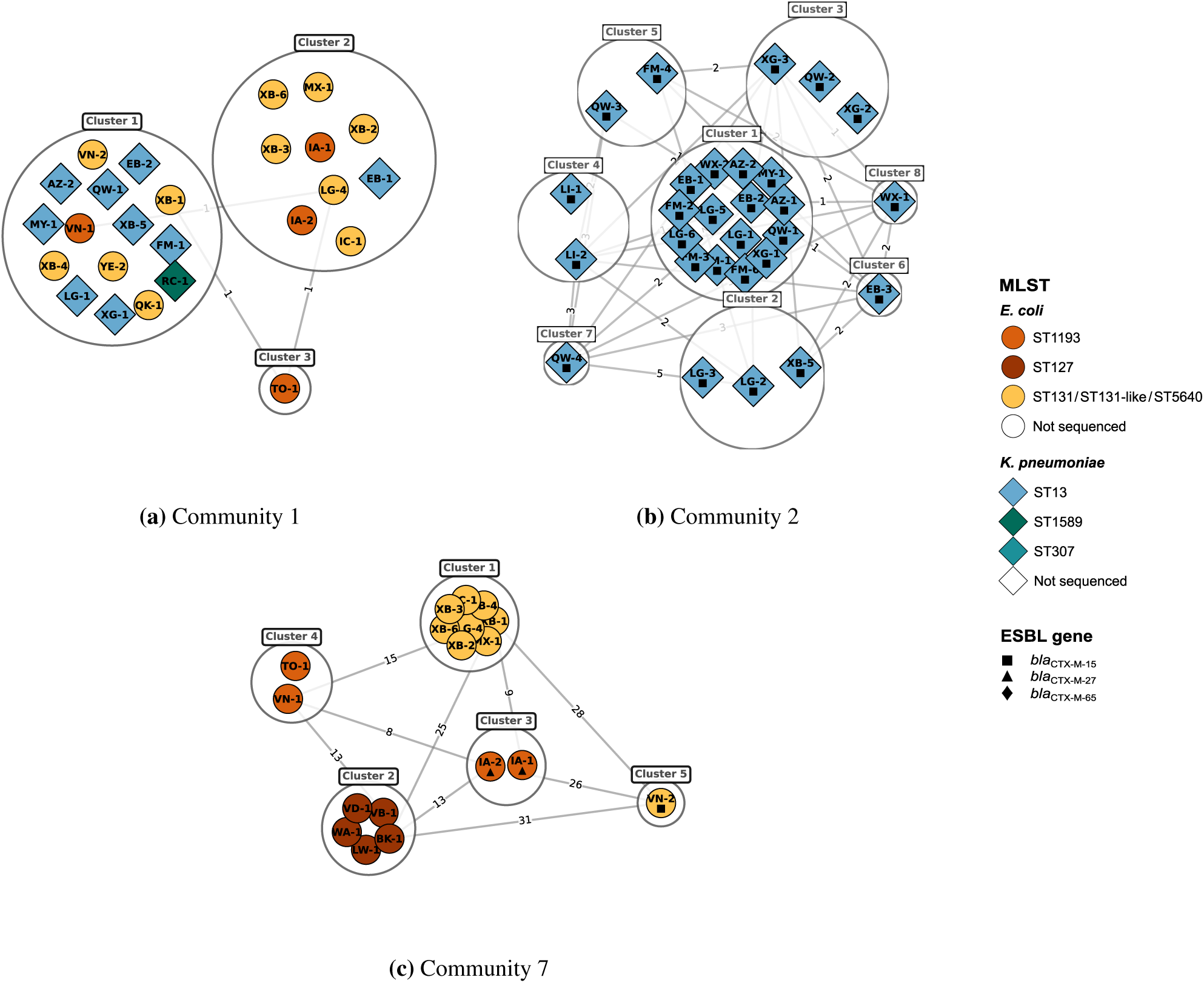
Network plots of selected plasmid communities. Each node represents a plasmid, labelled with the patient ID and sequenced-sample number it came from. Edges represent DCJ distance. For clarity, the network is compressed by clustering isolates with zero pairwise DCJ distance. Only plasmids that circularised during assembly are included. Plasmids bearing a *bla*_CTX-M_ allele are shown with a black symbol inside the node. (a) Community 1 shows Col-type plasmids, predicted to be mobilisable by MOB-suite. These are multi-species and multi-sequence-type and do not carry resistance genes. (b) Community 2 shows a large community of plasmids all belonging to *K. pneumoniae* ST13, all carrying the *bla*_CTX-M-15_ gene. These are predicted to be non-mobilisable by MOB-suite. They are large (around 200 kb) and have replicon type IncFIB(K). (c) Community 7 shows a community of plasmids belonging to isolates from three different *E. coli* sequence types, bearing two different *bla*_CTX-M_ gene variants, all with IncF replicons identified. Each CTX-M variant was confined to a single patient, and the corresponding 0-DCJ clusters were separated from all other 0-DCJ clusters by at least 8 DCJ rearrangements.

In the next largest plasmid community containing at least one *bla*_CTX-M_-positive plasmid, C7, 3/18 plasmids contained a *bla*_CTX-M_ gene (*bla*_CTX-M-15_, n=1; *bla*_CTX-M-65_, n=2), while 15/18 did not (Figure 3c). The remaining three plasmid communities each consisted solely of plasmids from single isolates or pairs of isolates from a single patient, indicating no evidence of wider plasmid dissemination.

Interestingly, several of the non-*bla*_CTX-M_-carrying plasmid communities (C1, C3, C4, C6, C9, and C10) contained members identified as not only being from either *K. pneumoniae* or *E. coli* but also originating from multiple MLST sequence types (Figure 3a). Most of these plasmid communities contained Col-type replicons and were predicted to be mobilisable via MOBP or MOBQ relaxases. These observations are consistent with either frequent plasmid exchange between species and MLST sequence types in patients or the unit, or the presence of a small number of conserved plasmids circulating across both species. As Col-type plasmids from external databases also clustered closely with our Col-type plasmids when included in the clustering analysis, there is insufficient evidence to support recent plasmid transfer within the timeframe of our dataset (Figure S5).

### Phylogenetic and single-linkage SNP clustering analysis supports presence of multiple clonal bacterial transmissions between patients

Phylogenetic analysis was performed on MLST-defined groups containing more than three isolates: *K. pneumoniae* ST13 and *E. coli* ST131, ST127, and ST1193. Isolates classified as ST131-like, and ST5640 isolates were also included in the ST131 group (Table S1). Phylogenetic maximum-likelihood trees were produced and annotated with corresponding plasmid community data (Figure 4).

**Figure 4:**
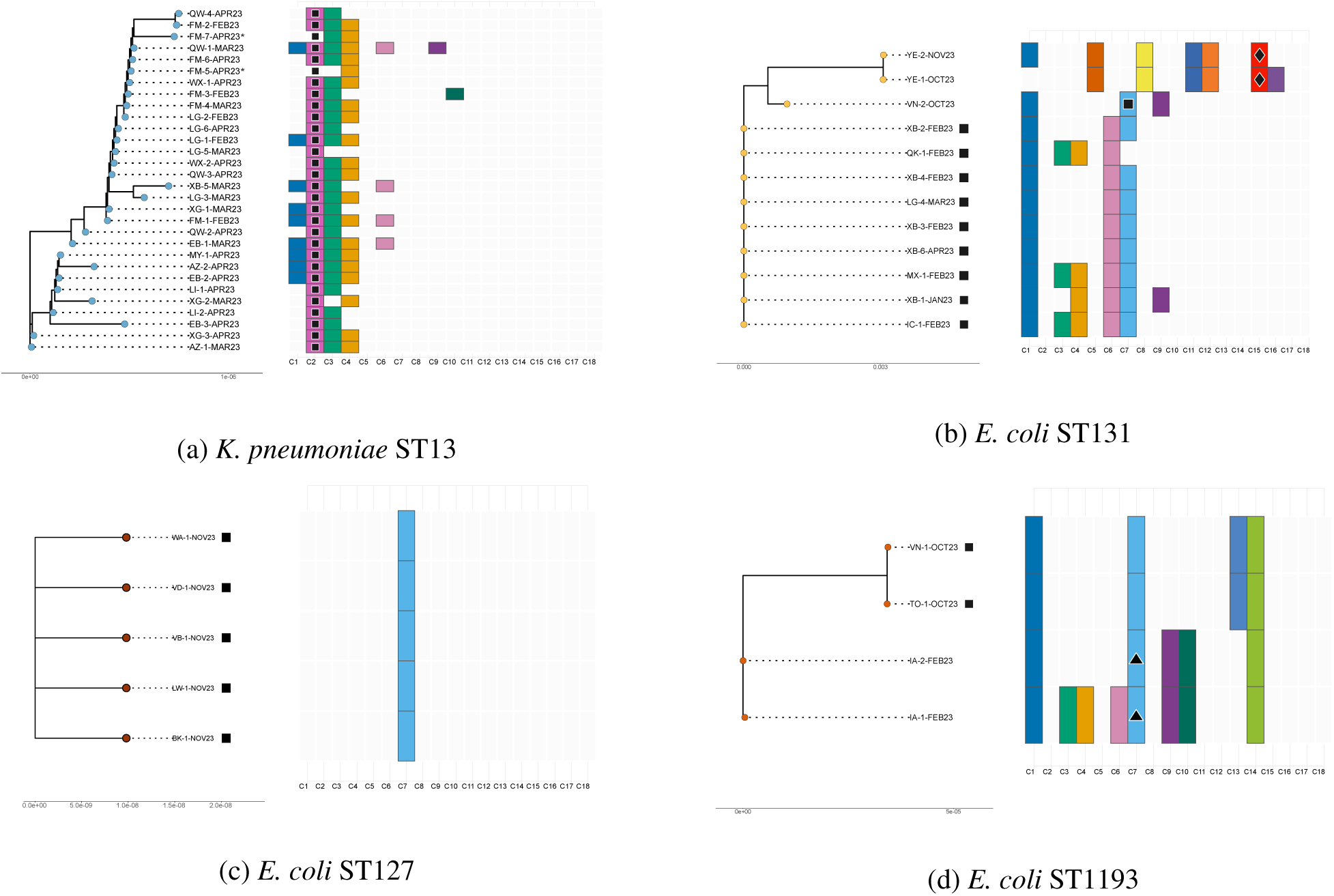
Phylogenetic trees with corresponding plasmid communities. Phylogenetic trees for each MLST group with annotation panels for the corresponding plasmids of each isolate. (a) *K. pneumoniae* ST13. (b) *E. coli* ST131/ST131-like. (c) *E. coli* ST127. (d) *E. coli* ST1193. (*) The pling analysis only included circular plasmids—FM-5 and FM-7 both contained the CTX-M-bearing plasmid. These isolates are estimated to be in this pling community based on contig replicon type presence. They were not included in the pling analysis due to their poor assembly (Figure S7). ARG symbols: ▪ bla_CTX−M−15_, ♦ bla_CTX−M−65_, and ▴ bla_CTX−M−27_.

The ST13 phylogenetic tree was tightly clustered, and all isolates had relatively uniform plasmid content, with most harbouring plasmids from C2 (*bla*_CTX-M-15_-IncFIB(K)(pCAV1099-114)), C3 (Col440I) or C4 (Col440II), with a few carrying other plasmids. The plasmid communities of the ST131 and ST1193 isolates were less similar to one another and there was greater plasmid variation between isolates. The ST127 isolates showed no diversity in both host phylogenetic and plasmid composition; it contained a single multi-replicon plasmid (Col156, IncFIB-AP001918 and IncFII-29) belonging to C7. These observations motivated a higher-resolution analysis of host genomic relatedness using SNP-based clustering.

Single-linkage clustering was therefore performed on the same isolate groupings used above. All single-linkage analysis was run with a SNP threshold of ≤ 7. Each MLST group contained exactly one cluster comprising isolates from more than one patient, with the remaining clusters containing isolates from a single patient only (Table S2 and Figures S14 and S15). All ST13 isolates formed a single large cluster (n=30), comprising isolates from ten different patients. These isolates all carried the *bla*_CTX-M-15_ variant on a plasmid. For the ST131 group, all isolates collected between January and April 2023 clustered together (n=9/12); these were collected from five different patients. These nine isolates all carried the *bla*_CTX-M-15_ variant on the chromosome. The other three isolates (collected in October and November) were singletons. Two of these isolates originated from the same patient (YE), both with *bla*_CTX-M-65_ on a plasmid, and differed by 27 SNPs. All five ST127 isolates from five different patients clustered together with zero pairwise SNP distance, consistent with a clonal lineage shared across patients. The four ST1193 isolates formed two pairs: one pair from a single patient and the other pair spanning two patients, indicating only one possible inter-patient transmission link within this sequence type. Altogether, the four multi-patient clusters identified in the single-linkage analysis suggest four distinct transmission clusters. In total, a minimum of 18 inter-patient transmission links were inferred based on cluster sizes (nine for ST13, eight for ST131, four for ST127, and one for ST1193); 20/23 (87%) of patients were involved in transmission links among sequenced isolates.

## Discussion

Plasmids play a central role in the horizontal dissemination of ESBL genes, with certain incompatibility groups contributing disproportionately to their global expansion [26]. Amoung these, IncF-type plasmids, particularly those carrying *bla*_CTX-M-15_, are widely found in Enterobacterales and reflect this capability for dissemination [27]. Because ESBL-encoding plasmids are capable of inter-species transfer [24], the identification of ESBL-producing isolates from multiple Enterobacterales species in the Oxfordshire neonatal ward in 2023 raised the possibility that a shared mobilisable ESBL-encoding plasmid was the source of the outbreak. However, long-read genetic sequencing found no evidence of a plasmid shared across species or between sequence types that could have driven ESBL dissemination within the ward. Instead, *bla*_CTX-M_–mediated resistance reflected multiple independent introductions into the ward, while highlighting at least four clonal transmission clusters involving 20/23 patients evaluated.

A common *bla*_CTX-M_-carrying plasmid with the replicon type IncFIB(K)(pCAV1099-114) was identified; however, this plasmid was confined to *K. pneumoniae* ST13 isolates and was predicted to be non-mobilisable. Together with the phylogenetic and SNP-based analyses, these findings support clonal expansion of the ST13 lineage rather than horizontal transfer of this *bla*_CTX-M_-bearing plasmid.

Phylogenetic analysis identified four clonal inter-patient clusters, indicating detectable patient-to-patient transmission among the IPC-flagged ESBL cases that underwent sequencing. The largest clonal cluster comprised ten patients, while the remaining clusters comprised five, five, and two patients. Because genomic analyses were limited to the available sequenced isolates, these findings likely represent a conservative estimate of transmission. Overall, the results are consistent with in-ward spread and suggest that continuing to strengthen standard transmission-based IPC measures may be warranted.

IncF plasmids are widely reported as being conjugative or mobilisable and are frequently associated with carriage of *bla*_CTX-M_ genes [28]. In our dataset, all large (c. 200 kb) ST13-associated IncFIB(K) plasmids were classified as non-mobilisable by MOB-suite, with no detectable relaxase or MPF genes identified. This differs from typical F-type plasmid backbones, which ordinarily include a *tra* locus encoding transfer-associated functions [29]. The restriction of this IncFIB(K) plasmid to ST13 in our dataset is therefore notable and may reflect the ancestral loss of transfer-associated genes, resulting in a non-mobilisable derivative in this lineage.

In contrast, multiple smaller, inter-species, non-*bla*_CTX-M_-bearing plasmid clusters were predicted to be mobilisable, including Col-type replicons such as Col156. Their presence across different species could suggest the possibility of recent interspecies horizontal transfer within our dataset. However, comparison with globally distributed Col-type plasmid sequences obtained from the PLSDB database and GenBank demonstrated similarly high sequence conservation, indicating that these plasmids are highly stable and widely disseminated, reducing the likelihood of any recent inter-species transfer events within our dataset (Supplementary S5). There can be difficulty distinguishing recent plasmid dissemination from a widely disseminated and stable plasmid, where the usual SNP-based approaches used for clonal comparison may not be applicable due to the high rate of recombination events commonly seen in plasmids; therefore, the inclusion of global data should be considered in these structural clustering analyses to provide additional context. Although none of the single-replicon Col-type plasmids identified in this study carried *bla*_CTX-M_ genes, previous reports have shown that clinically significant resistance determinants can integrate into such backbones. For example, the integration of *bla*_OXA-48_ into a Col156 plasmid was associated with a carbapenemase-producing outbreak in New Zealand [30]. Such findings highlight the importance of considering these plasmids in ongoing genomic surveillance.

Several observations are worth noting. First, two ST1193 isolates from the same patient were 29 SNPs apart. Given the short sampling interval and the low likelihood of such divergence over this period, this difference is most plausibly attributable to sequencing or mapping artifacts, although within-host diversity cannot be excluded. This observation suggests that more stringent variant masking or more conservative single-linkage thresholds could be applied; however, there was no evidence of *bla*_CTX-M_ transmission beyond this patient, and this finding therefore does not affect the overall interpretation of transmission dynamics. Second, the combined sample chosen for FM-7, an isolate sequenced twice to validate the phylogenetic analysis and also one of the isolates with a poorly assembled *bla*_CTX-M_-carrying plasmid had two *bla*_CTX-M-15_ genes found in the same sample. However, since the plasmid had not properly assembled, it was decided that it was unknown whether or not this was a genuine repeat due to a duplication of a mobile genetic error, or a sequencing error. Third, although this does not hugely change the main findings of our study, the lone *E. coli* isolate among multiple *E. coli* isolates from patient LG and lone *K. pneumoniae* isolate from patient XB among multiple *E. coli* isolates should be highlighted, and the possibility of a sample, library or other switch must be considered, especially since samples were collected on the same day.

Our study has several limitations. Due to resource constraints, only a subset of ESBL-positive isolates underwent sequencing, and ESBL-negative isolates were not included. A broader sampling framework may have provided greater resolution to assess plasmid dynamics, other mobile genetic element movement (such as transposons etc.), and potential transmission pathways within the ward. Although our findings are most consistent with multiple independent introductions, alternative mechanisms of resistance gene dissemination remain possible. More comprehensive characterisation of mobile genetic elements, together with expanded genomic and environmental sampling, would help to further resolve transmission dynamics in future work. Technical and analytical limitations should also be considered. Although Oxford Nanopore Technologies sequencing accuracy has improved substantially, complete resolution of all plasmids remains challenging, particularly across repetitive regions. Although heavily fragmented genomes were re-sequenced to improve assembly quality, several assemblies remained incomplete, particularly across repetitive regions, which prevented us resolving all the plasmids in the dataset. Plasmid clustering with Pling relies on DCJ distance thresholds for inferring relatedness but biologically validated cut-offs for transmission inference are not yet standardised. In addition, the tools used to identify replicon types, relaxases, and MPF systems depend on reference databases, and divergent or uncharacterised genes may not have been detected, meaning we may have potentially underestimated plasmid mobility or other plasmid features.

The diversity of *bla*_CTX-M_ variants observed in the OUH NICU in 2023 suggests multiple independent introductions of ESBL-producing bacteria rather than horizontal dissemination mediated by a common plasmid. However, the identification of several clonal transmission clusters demonstrates that in-ward spread was likely still occurring, which was addressed by IPC. Although plasmid-driven inter-species outbreaks have been reported on multiple occasions for carbapenemase genes, and remain biologically plausible for ESBL genes, our findings indicate that this mechanism was not the dominant driver in this putative outbreak. Importantly, the analytical pipeline developed in this study can now be applied to future putative ESBL outbreaks. The ability to distinguish between multiple introductions, clonal transmission, and horizontal dissemination of ESBL genes, including through plasmids and other mobile genetic elements, is important to guide IPC strategies, particularly in high-turnover neonatal settings. The identification of multi-species Enterobacterales clusters should therefore be investigated carefully. Genomic analysis can be used to clarify the underlying mechanism(s). Long-read sequencing enables resolution of these different mechanisms and is therefore likely to enhance future surveillance frameworks. Together, these findings highlight the value of integrating genomic approaches into IPC practice to refine outbreak assessment and guide proportionate, targeted interventions.

## Methods

### Electronic healthcare data extraction

We used data from the Infections in Oxfordshire Research Database (IORD) [31], which includes de-identified medical records from four hospitals comprising the Oxford University Hospitals (OUH), results from samples sent from the community and hospital for testing at the OUH laboratories, and hospital prescribing. These hospitals serve a population of approximately 755,000 individuals. To ascertain baseline admission, testing and culture-positivity rates on the unit, access to the relevant data extracts from IORD was secured, thereby providing the epidemiological context of the outbreak. Ethical permissions for the sequencing and outbreak evaluation are covered as part of a wider Research Ethics Committee (REC) application (REC ref: 17/LO/1420).

### Sample collection

The study was undertaken using clinical isolates collected from neonates in the Oxford University Hospitals (OUH) NHS Foundation Trust, Oxford UK. Given ESBL-associated infections had previously been detected on the unit, and in the context of a rectal screening programme including admission, weekly and discharge swabbing, the occurrence of multiple ESBL-positive isolates among neonates with poor clinical outcomes prompted investigation by the IPC team, and 53 third-generation-cephalosporin-resistant isolates that had been phenotypically flagged as ESBLs by routine diagnostic methods (32 *K. pneumoniae* and 21 *E. coli*) were cultured from 23 neonatal inpatients (January 2023 to November 2023).

### ONT sequencing

Bacteria were sub-cultured from frozen stocks on Columbia Blood Agar (CBA) and incubated at 37°C overnight. 10 µL loops of culture underwent DNA extraction using the QIAGEN Genomic Tip extraction kit. DNA was stored at 4°C until ready for sequencing. Sequencing was performed on an Oxford Nanopore Technologies GridION, using an R10.4.1 flow cell and the Rapid Barcoding kit (RBK114.96). The 53 samples were initially sequenced across four flowcells: The 42 samples collected between 02.01.23 and 23.04.23 were split across three flow cells, with the remaining 11 samples collected between 07.10.23 and 20.11.23, sequenced on the fourth. Of the 53 isolates sequenced, 46 passed quality control (QC) first time and seven passed following resequencing. Samples were classified as ‘poor quality’ and failed QC if they did not assemble well. The QC metric was decided using both visual inspection of the bandage plots and with the inter-quartile range of the number of contigs per sample. A fifth flowcell accommodated these repeats along with a further seven repeats of isolates that passed QC which were selected for validating our phylogenetic analysis, making a total of 14 samples (Figure S2). Basecalling was performed with Dorado v0.8.1 SUP. The first batch was processed using the model dna_r10.4.1_e8.2_400bps_sup@v4.1.0, while the remaining batches were basecalled with dna_r10.4.1_e8.2_400bps_sup@v5.0.0, selected according to their respective sequencing sampling frequencies [32] (Figure S4).

### Genome assembly and general analysis

An automated Nextflow pipeline (v23.04.1) was developed to assemble the bacterial genomes (Figure S3) and is publicly available [33]. It takes metadata and raw reads as input and passes the reads through an assembly process [34–36]. Scripts within Nextflow were executed using Python v3.13.3 and R v4.3.3. Multiple quality control steps were performed using SeqKit v2.5.1, and reads were filtered using NanoQ v0.10.0 [37, 38]. Reads were subsampled to 150× coverage using Rasusa v0.7.1 with a fixed random seed (–s 35) [39]. Genomes were assembled with Flye v2.9.2 [34], visualised with Bandage v0.8.1 [40], and circular contigs were rotated using dnaapler v0.7.0 [36]. Assemblies were then polished with Medaka v1.8.0, using models r1041_e82_400bps_sup_v4.1.0_model and r1041_e82_400bps_bacterial_methylation_model for the first and remaining batches, respectively [35].

Following assembly, downstream analyses included multilocus sequence typing using MLST v2.23.0 and antimicrobial resistance gene identification using AMRFinderPlus v3.11.26.

Outside of the Nextflow pipeline, preprocessing and postprocessing scripts were used for data manipulation, analysis and plotting [41].

### Plasmid clustering

Pling v2.0.0 [42] was applied to cluster similar plasmids using the Pling containment distance and then DJC-distance; this was run with the sourmash containment setting and default parameters. Pling analised 73 plasmids, defined as contigs with circular topology and a length under 1,000,000 base pairs. For each plasmid community, network plots with added metadata (species, MLST, patient labels, *bla*_CTX-M_ variants) were drawn in Python using Matplotlib v3.10.3 and NetworkX v3.2.1 [43, 44]. Nodes were compressed where DCJ distance was zero, forming further clusters, simplifying plots with edge removal. The average edge length between each 0-DCJ cluster was taken and rounded to the nearest whole number, further decluttering the plots. Additionally, PlasmidFinder v2.1.6 with database v2.1 was used to identify plasmid replicons in each contig, and MOB-suite v3.1.8 was used to detect relaxase genes, MPF systems, and to predict plasmid mobility [45, 46].

### Phylogenetic analysis and single-linkage SNP clustering

A downstream phylogenetic analysis subpipeline was developed in Nextflow (Figure S4).

For each MLST group containing more than three isolates, a reference genome was selected from assemblies within the dataset based on the earliest collection date with high assembly quality. The selected references were XB-2 for ST131, VB-1 for ST127, FM-1 for ST13, and IA-1 for ST1193.

Reads were mapped to their respective reference genomes using Minimap2 v2.28 with default settings [47], and variant calling was performed using Clair3 v1.0.10 with default parameters. Clair3 was selected because deep learning-based variant callers have been shown to outperform alternative methods for bacterial ONT sequencing data [48, 49]. A parameter sweep was conducted to optimise variant masking thresholds, balancing the reduction of false positives with the retention of sensitivity, which is particularly important for outbreak datasets where genetic distances between isolates are small (Figure S6). The final masking parameters were a minimum read depth of 20×, a minimum variant quality score of 5, and a minimum allele frequency threshold of 0.9. Variant call format (VCF) file masking was performed using SAMtools v1.15.1 and BCFtools v1.17.

For each sample, consensus sequences were generated using BCFtools v1.20 [50]. Phylogenetic trees were inferred using IQ-TREE v2.3.5 with default settings [51], and visualised in R v4.4.1 [52] using ggplot2 v3.5.1, ggtree v4.1.1 and ggtreeExtra v1.16.0 [53, 54]. Figures were refined using Inkscape v1.4.

Pairwise SNP distances were calculated using SNP-dists v0.8.2. Single-linkage clustering was performed in R v4.3.3 using the package igraph v2.1.4 with a ≤7 SNP threshold. This threshold is broadly consistent with estimated within-host evolutionary rates of approximately 3–4 SNPs per genome per year for *E. coli* and 1–4 SNPs per genome per year for *K. pneumoniae* [55]. A sensitivity analysis showed that varying the threshold from 5 to 20 SNPs did not alter cluster assignments.

## Acknowledgements

This work was funded by the National Institute for Health and Care Research (NIHR) Biomedical Research Centre (BRC), Oxford (NIHR203311), and the NIHR Health Protection Research Unit in Healthcare Associated Infections and Antimicrobial Resistance at Oxford University in partnership with the UK Health Security Agency (UKHSA) (NIHR207397). This work uses data provided by patients and collected by the UK’s National Health Service as part of their care and support. We thank all the people of Oxfordshire who contribute to the Infections in Oxfordshire Research (IORD) Database. Research Database Team: L Butcher, H Boseley, C Crichton, DW Crook, D Eyre, O Freeman, J Gearing (community), R Harrington, K Jeffery, M Landray, A Pal, TEA Peto, TP Quan, J Robinson (community), J Sellors, B Shine, AS Walker, D Waller. Patient and Public Panel: G Blower, C Mancey, P McLoughlin, B Nichols. The views expressed are those of the authors and not necessarily those of the NHS, the NIHR, the Department of Health and Social Care or the UK Health Security Agency. Computation used the Oxford Biomedical Research Computing (BMRC) facility, a joint development between the Wellcome Centre for Human Genetics and the Big Data Institute supported by Health Data Research UK and the NIHR Oxford Biomedical Research Centre. The views expressed are those of the authors and not necessarily those of the NHS, the NIHR, the Department of Health and Social Care or UKHSA. For the purpose of Open Access, the author has applied a CC BY public copyright licence to any Author Accepted Manuscript (AAM) version arising from this submission.

## Author contributions

M.J.P. designed and implemented the bioinformatics pipelines, performed the bioinformatics and statistical analyses, generated all figures, and contributed to study design and data interpretation. K.H. performed all of the sequencing. K.H., N.St., and B.Y. extracted clinical metadata. J.C. extracted the relevant electronic healthcare records. S.O., K.J., S.P. performed patient sample collection and L.Ba. selected samples of interest and described isolation rates on the ward. K.C., D.E., N.St. originally supervised the study; N.Sa., P.W.F., P.B. later supervised the study. M.J.P. wrote the original manuscript, and P.W.F., N.Sa., P.B. revised it. N.St. acquired funding. All the authors reviewed the manuscript.

## Data availability

The sequencing data that support the findings of this study have been deposited in the European Nucleotide Archive (ENA) under BioProject accession PRJEB112737 (study accession ERP193205) and will be made publicly available upon publication.

