## Supplementary for "Genomic Epidemiology of Multi-Modal ESBL Gene Transmission among Enterobacterales in a Neonatal Unit"

**Figure S1: ESBL-positive *E. coli* and *K. pneumoniae* isolates collected from the Oxford NICU throughout 2023, as recorded in IORD**

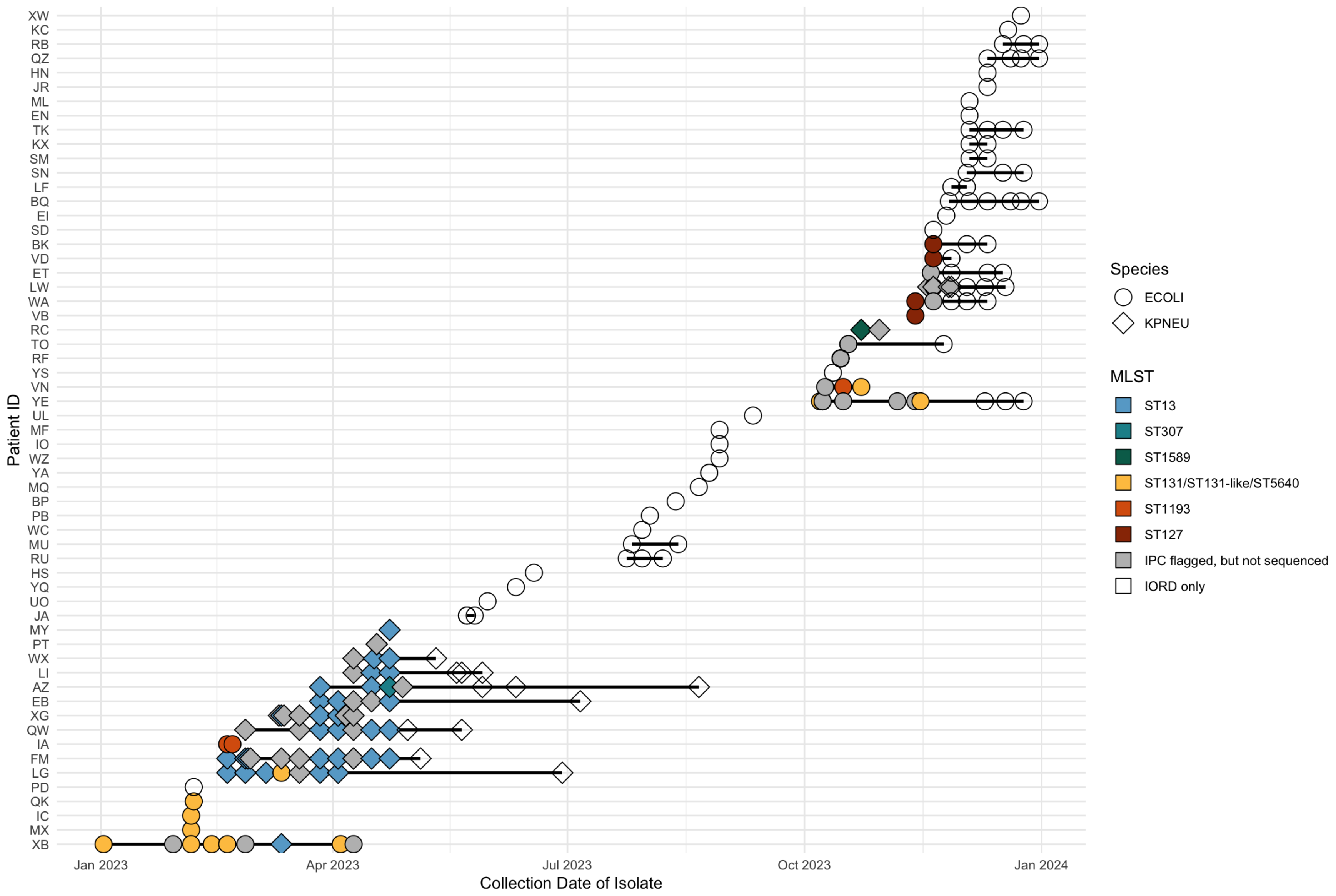

Time series plot of ESBL-positive isolates. Point shape indicates species. Isolates flagged by IPC for further investigation because of increased concern regarding ESBL-positive isolates in the ward are coloured according to MLST sequence type if sequenced, or grey if highlighted but not selected for sequencing. All other collected isolates that were not highlighted by IPC are shown as transparent points.

#### Figure S2: Comparison of assembly outputs for selected replicate isolates

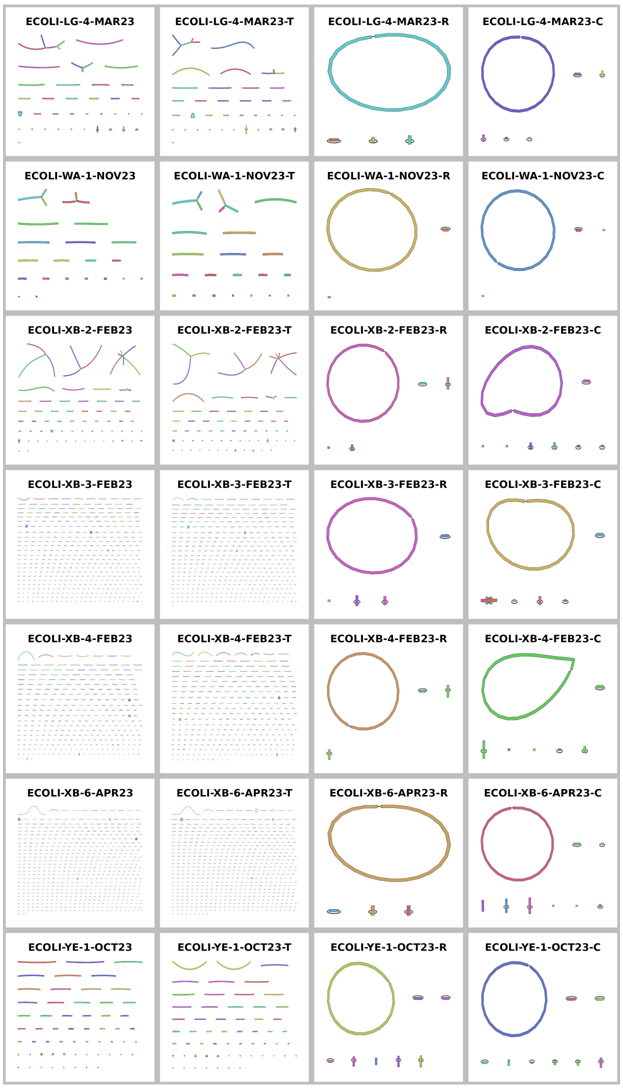

**(a)** Samples that originally failed quality control.

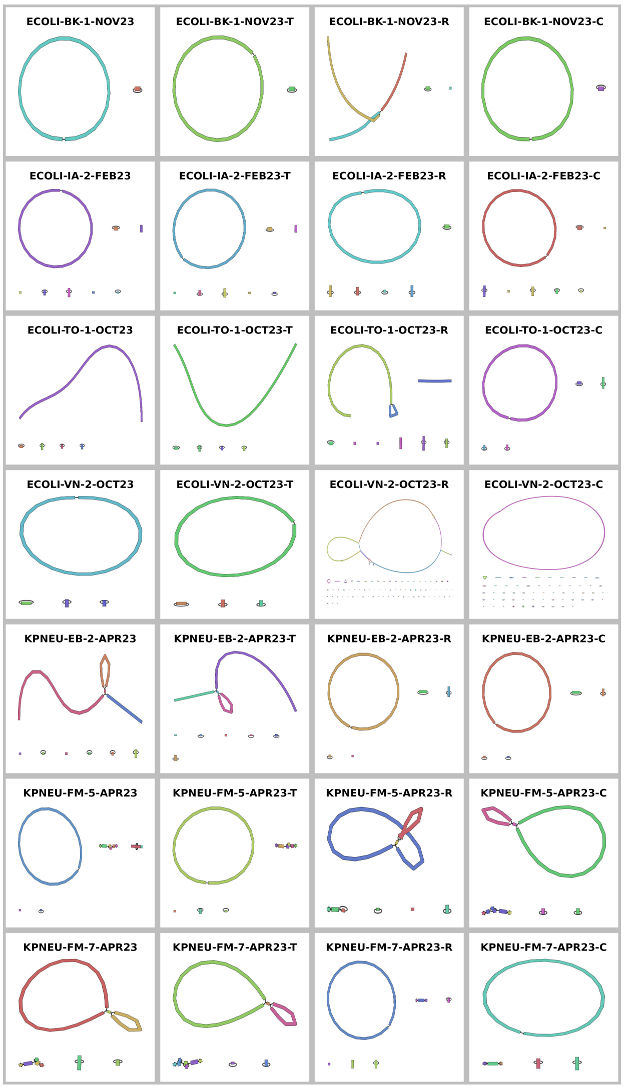

**(b)** Samples chosen as replicates for validation in phylogenetic analysis.

For each chosen isolate, four versions were compared: the original, a technical replicate (-T), a re-sequenced replicate (-R), and a combined sample (-C). Technical replicates were generated using the same reads as in the original sample and were included to assess the impact of non-deterministic steps in the assembly pipeline, e.g. the assembly tool Flye. Re-sequenced samples were generated either to improve assembly quality for samples that failed quality control, or for validation in the phylogenetic analysis. Combined samples were generated by concatenating the reads from both the original and re-sequenced samples to assess whether a larger number or reads and therefore increased read depth and coverage improved assembly quality.

**Figure S3: Overview of the main bioinformatics workflow, implemented in Nextflow.**

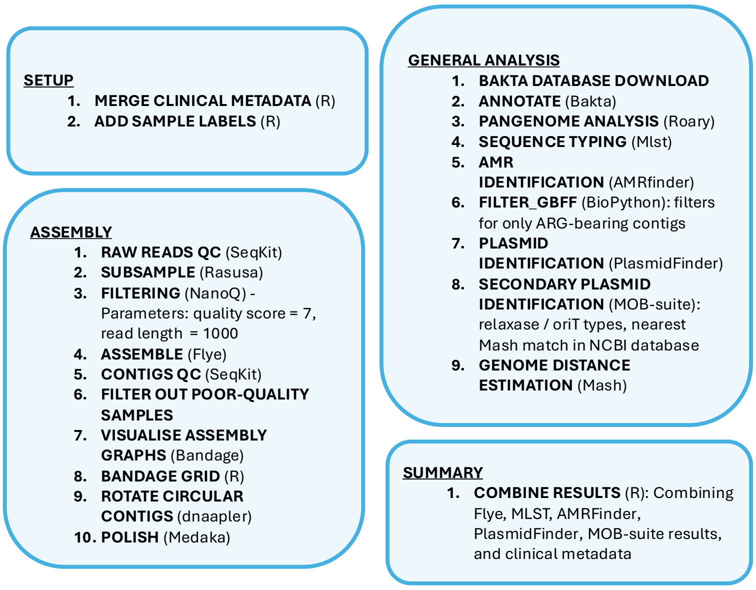

The pipeline consists of four stages: setup, genome assembly, general analysis, and summary. Sequencing data alongside corresponding metadata are processed to produce genotypic characterisation. Major processes and their functions are shown.

**Figure S4: Overview of the additional bioinformatics workflows that were implemented in Nextflow**

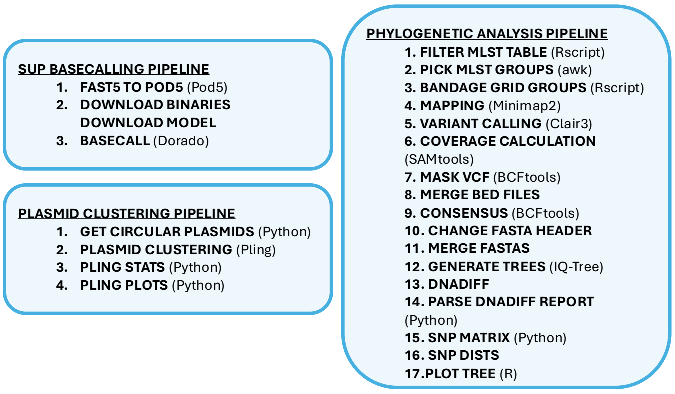

The SUP basecalling pipeline converts fast5 files to fastq files. This pipeline was created to improve the accuracy of the samples in our dataset, which were originally basecalled by the MinION machine with the HAC setting. The plasmid clustering pipeline was created separately to the main pipeline to facilitate a rerun with additional samples. This pipeline takes all circular contigs associated with plasmids and clusters then into plasmid communities. The graphs of these communities are then annotated with genomic and clinical metadata. The phylogenetic analysis pipeline groups samples based on MLST and creates a maximum liklihood tree for each group comprising more than three samples. This pipeline was also created separately from the main workflow to enable straightforward inclusion or exclusion of the validation samples.

**Figure S5: External Col-type plasmids added to the plasmid clustering analysis**

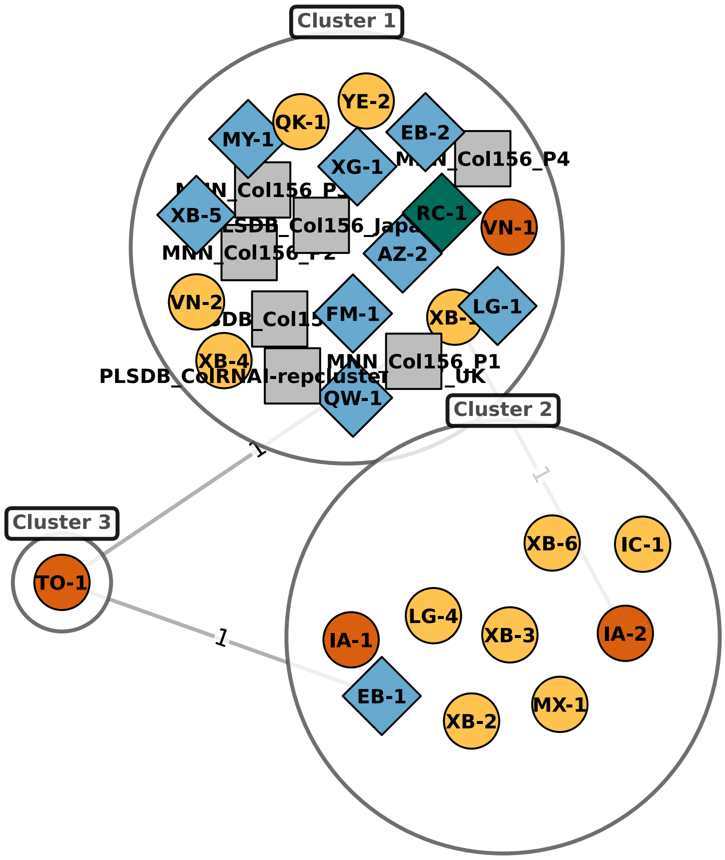

External plasmids are represented by grey squares. Since a few plasmid communities contained multiple species, were predicted "mobilizable", and had small DCJ distances between plasmids within each community, we investigated whether there was a transfer of these plasmids within our dataset or whether these are widely disseminated and stable Col plasmids. To do this, for each Col-type multi-species plasmid community, we found all mash-nearest-neighbour matches from MOB-suite and additionally chose plasmids of the same replicon type from the PLSDB database from Oxfordshire, elsewhere in the UK, and elsewhere in the world. We reran Pling, incorporating these external plasmids with our existing plasmid dataset to see if they would cluster with our plasmids. Many of the external plasmids clustered with our existing communities, and low DCJ distances were observed across many plasmids, including globally distributed ones, suggesting widespread dissemination and weakening the evidence for transfer within the study period

**Figure S6: Pairwise SNP distances between replicate isolates across masking parameter combinations**

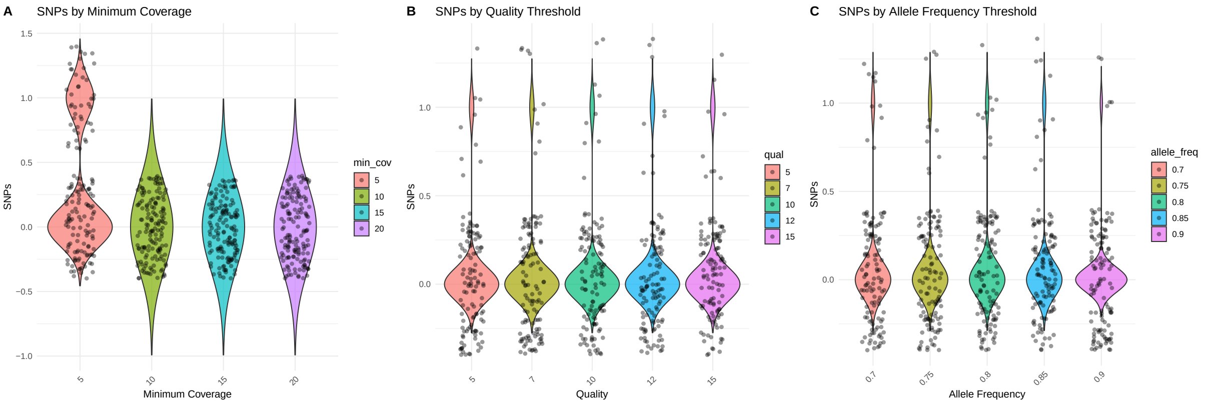

Six pairs of isolates that passed quality control were used for validation. The parameter set minimum coverage = 20, quality = 5, allele frequency = 0.9 was chosen.

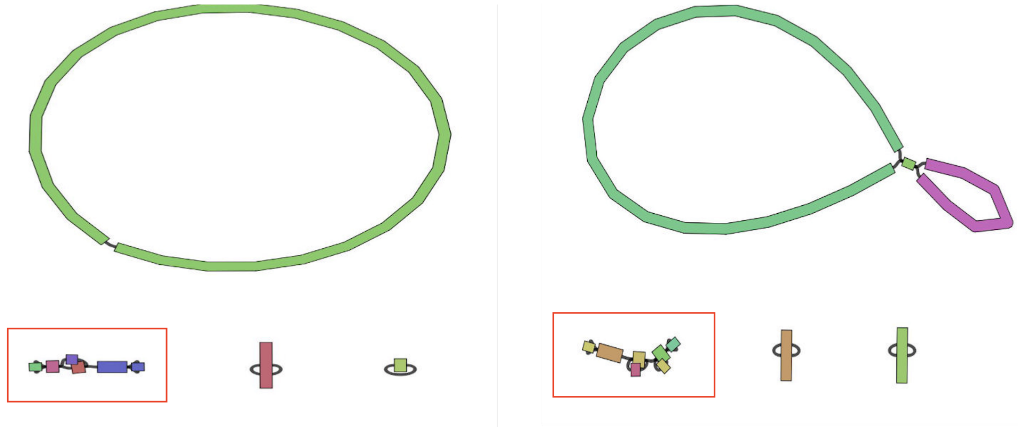

**(a)** KPNEU-FM-5-APR23.

**(b)** KPNEU-FM-7-APR23.

**Figure S7: Putative plasmid reconstruction comprising multiple contigs.** For isolates FM-5 and FM-7, both *K. pneumoniae* ST13, the bla$_{\mathrm{CTX-M-15}}$ gene was identified on a contig that could not be resolved into a complete circular plasmid sequence. Instead, the putative plasmid assemblies comprised multiple contigs, with the IncFIB(K) replicon and bla$_{\mathrm{CTX-M-15}}$ gene located on separate contigs within the reconstructed plasmid structure.

#### Figure S8: Plasmid clustering and features

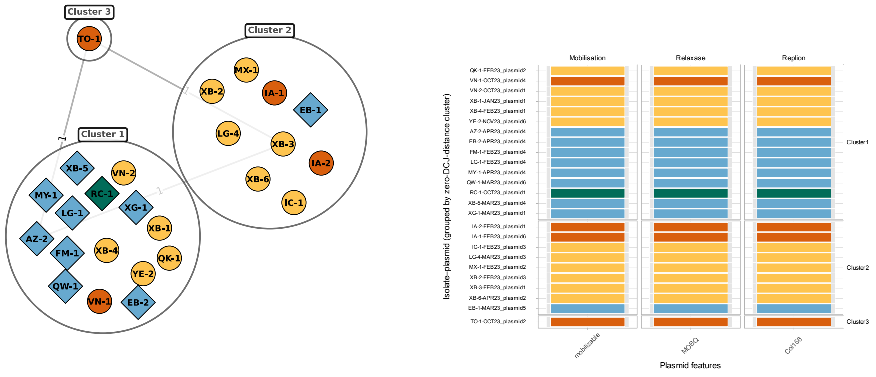

(**a**) Community 1

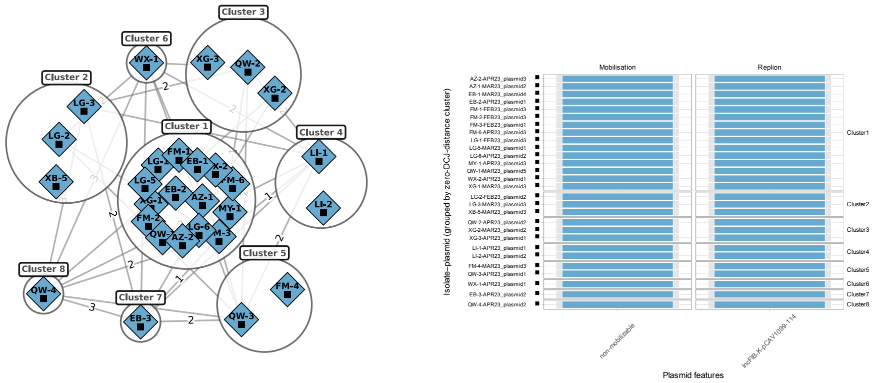

(**b**) Community 2

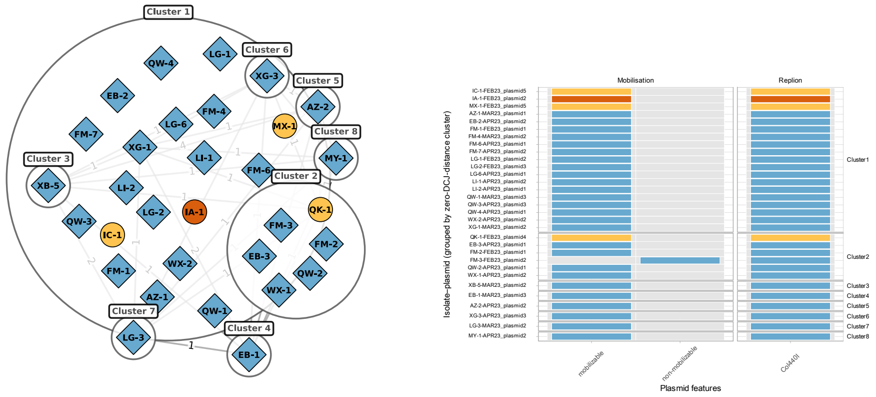

(**c**) Community 3

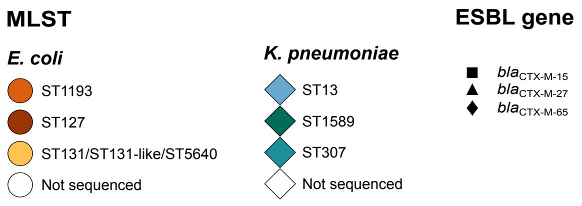

Left: plasmid network community plots. Right: corresponding plasmid feature plots.

**Figure S9: Plasmid clustering and features**

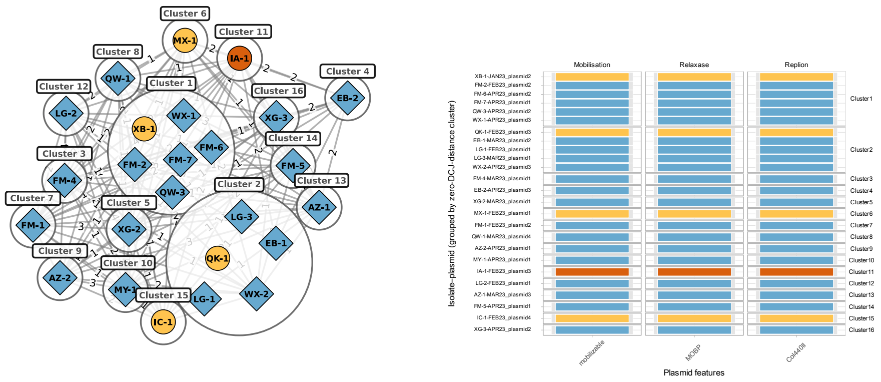

**(a)** Community 4

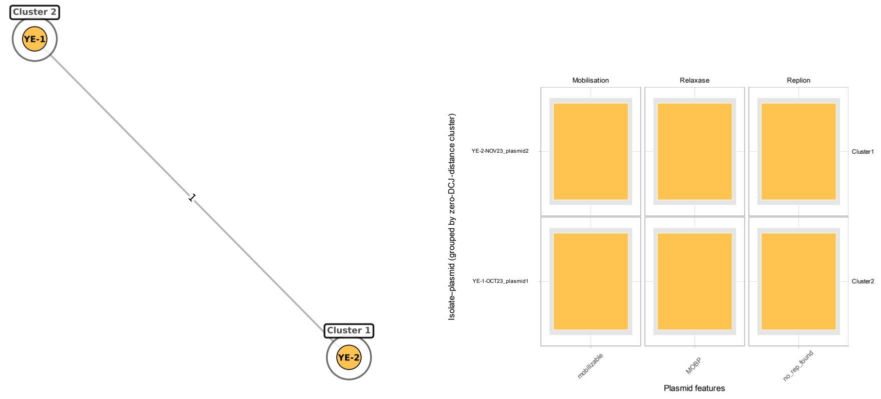

**(b)** Community 5

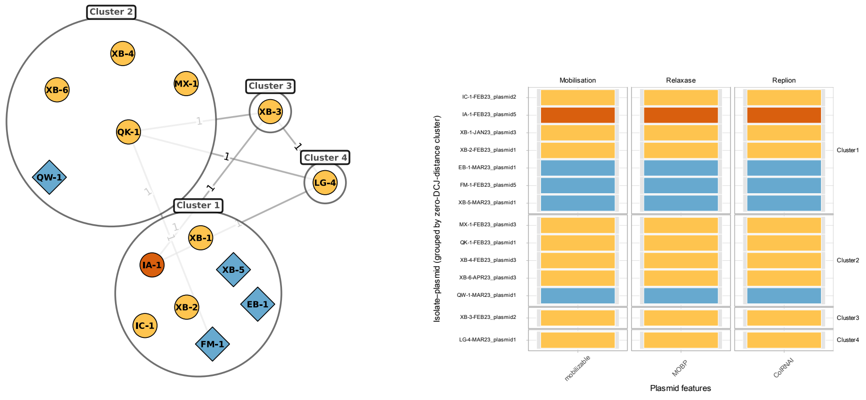

**(c)** Community 6

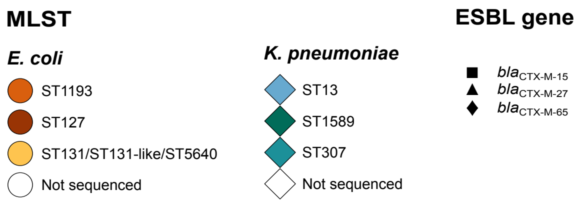

Left: plasmid network community plots. Right: corresponding plasmid feature plots.

**Figure S10: Plasmid clustering and features**

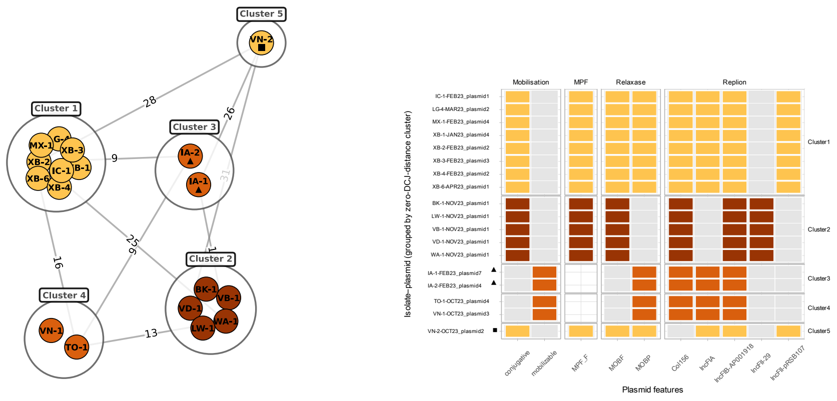

(**a**) Community 7

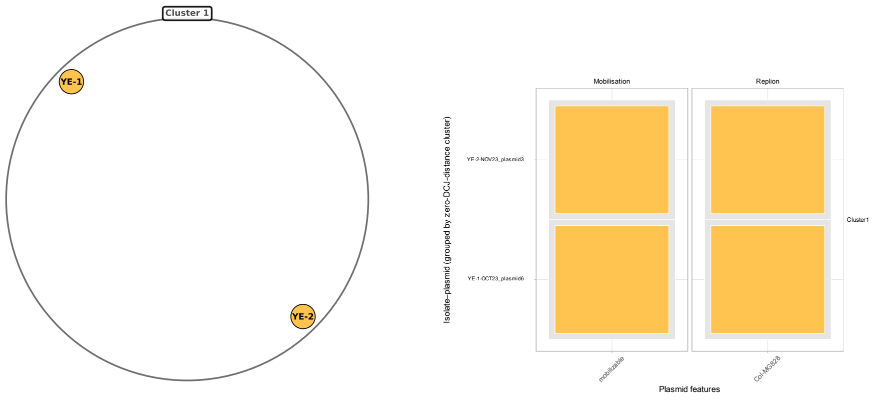

(**b**) Community 8

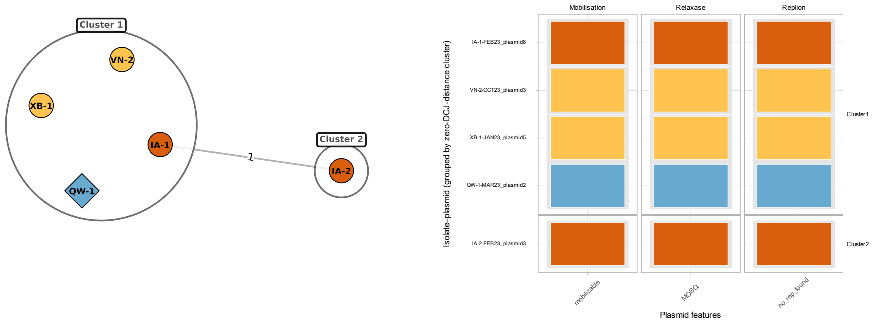

(**c**) Community 9

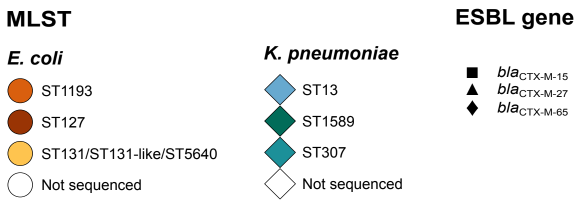

Left: plasmid network community plots. Right: corresponding plasmid feature plots.

#### Figure S11: Plasmid clustering and features

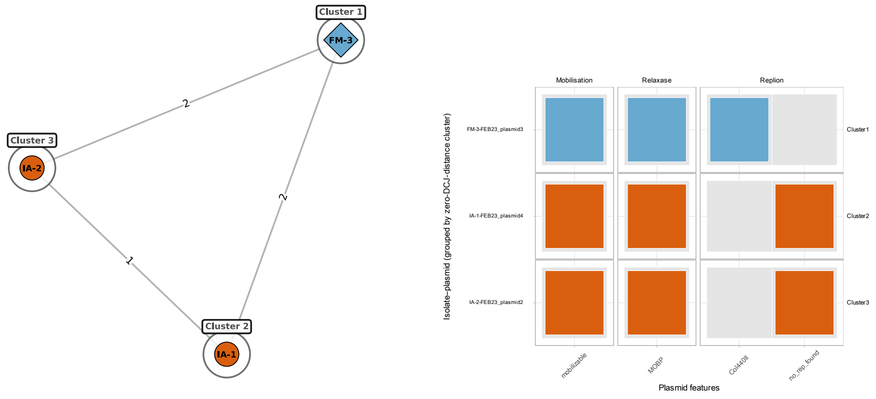

**(a)** Community 10

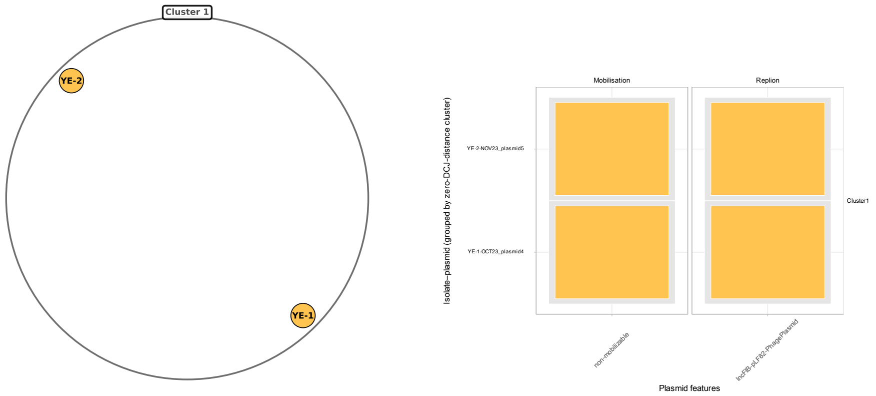

**(b)** Community 11

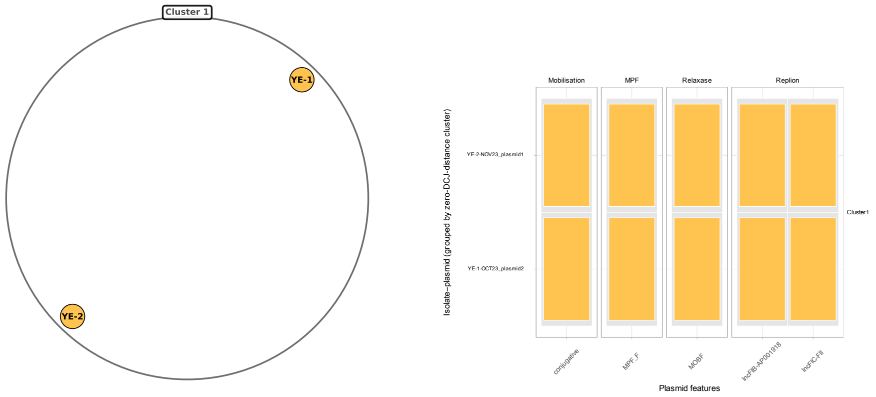

**(c)** Community 12

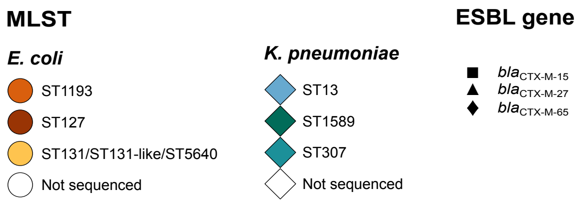

Left: plasmid network community plots. Right: corresponding plasmid feature plots.

### Figure S12: Plasmid clustering and features

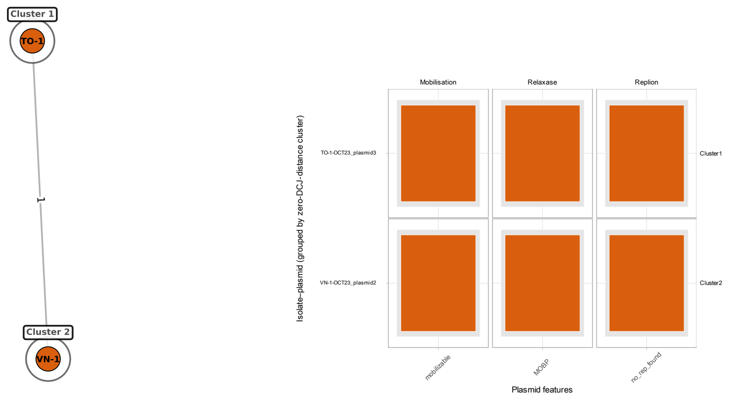

**(a)** Community 13

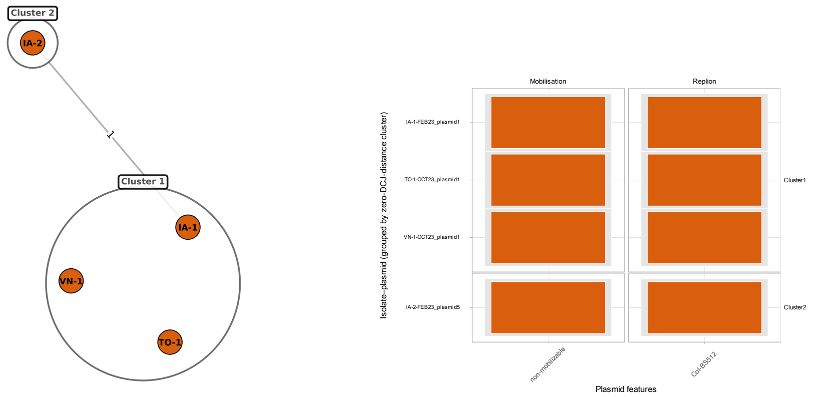

**(b)** Community 14

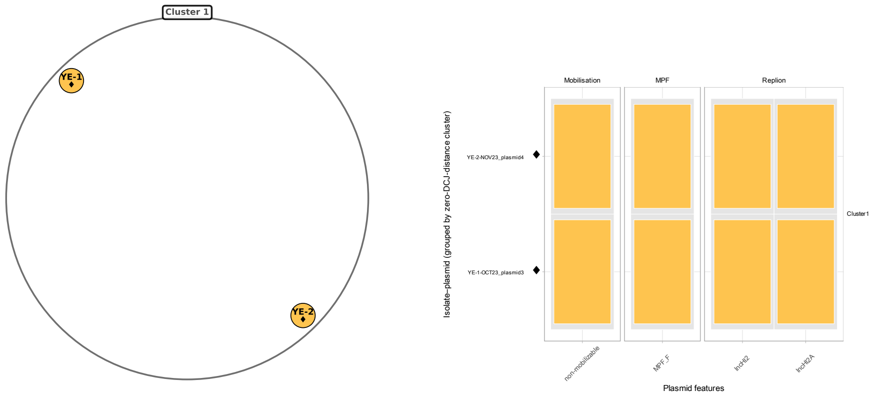

**(c)** Community 15

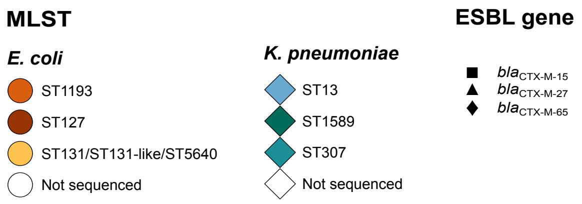

Left: plasmid network community plots. Right: corresponding plasmid feature plots.

#### Figure S13: Plasmid clustering and features

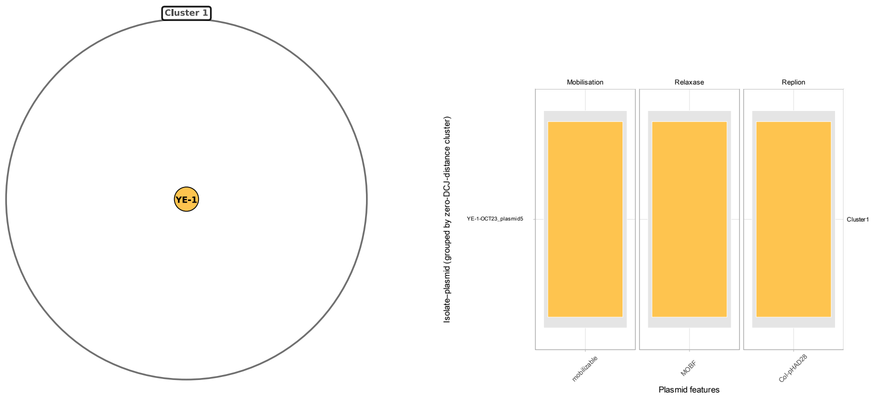

**(a)** Community 16

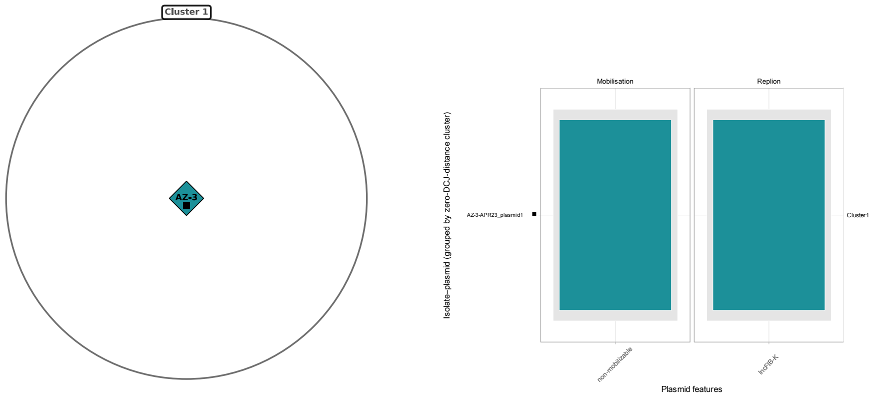

**(b)** Community 17

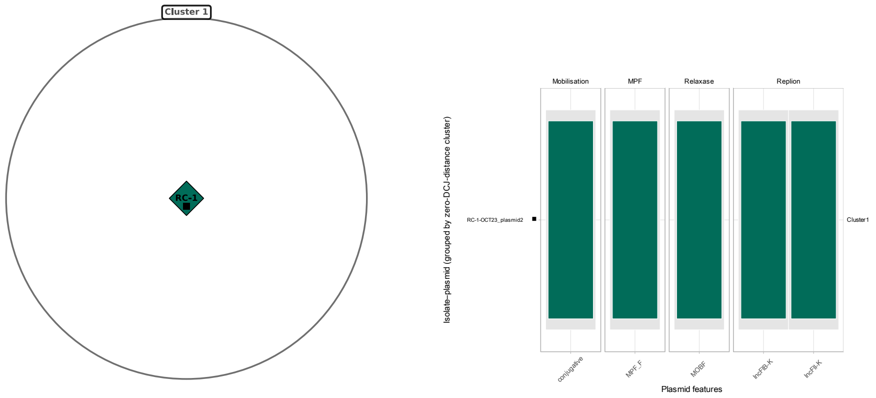

**(c)** Community 18

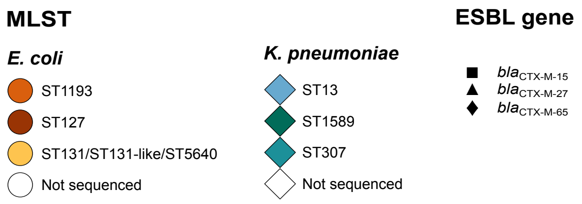

Left: plasmid network community plots. Right: corresponding plasmid feature plots.

**Figure S14: SNP distance between isolate chromosomes within MLST groups**

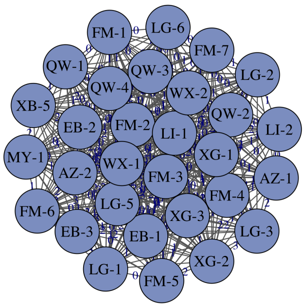

**(a)** *K. pneumoniae* ST13.

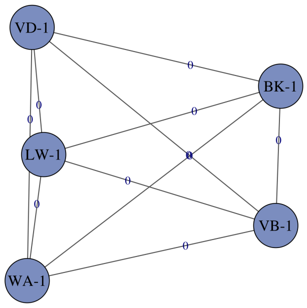

**(b)** *E. coli* ST127.

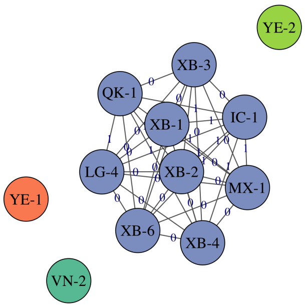

**(c)** *E. coli* ST131.

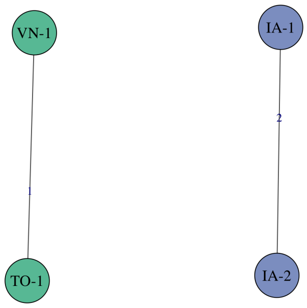

**(d)** *E. coli* ST1193.

Overall, eight clusters were identified, four of which (one from each MLST-type group) contained multiple patients. Cluster graph plots for the four main MLST groups. (a) ST13, containing a cluster of 30 samples from ten different patients. (b) ST127, showing a cluster of five samples from five different patients, all differing by zero SNPs, heavily implying clonal transmission between these patients. (c) ST131, showing a close cluster of nine samples from five patients suggesting a clonal outbreak. Three other samples in the same MLST group from two patients were not closely similar to the main clonal cluster. (d) ST1193, showing one case of potential transmission between patients, with two samples from two patients only one SNP difference between those samples. The other cluster of two samples was from the same patient.

#### Figure S15: SNP distance summaries between isolates

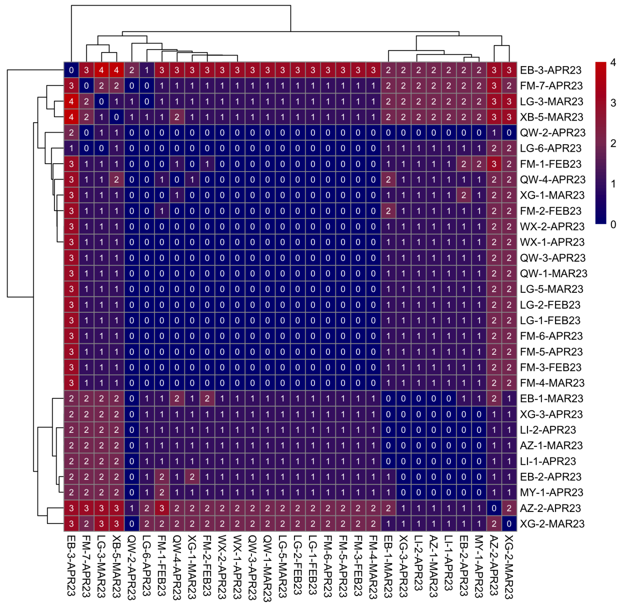

**(a)** *K. pneumoniae* ST13

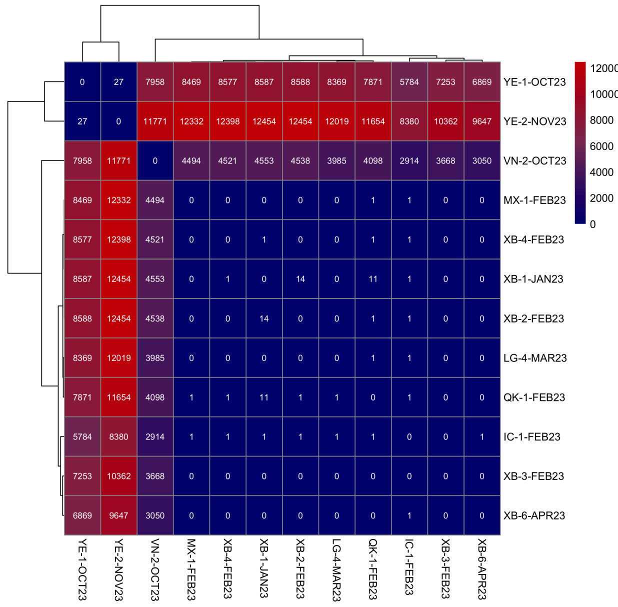

**(b)** *E. coli* ST131

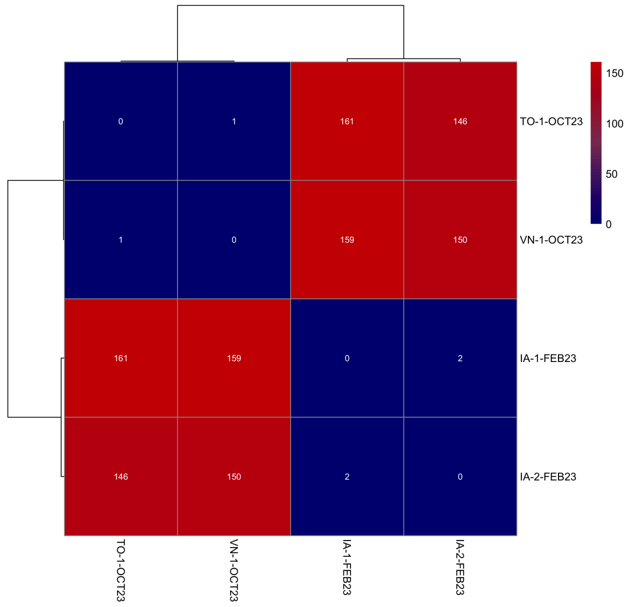

**(c)** *E. coli* ST1193

(a) *K. pneumoniae* ST13: all 30 isolates have less than or equal to four SNPs between them. (b) *E. coli* ST131: nine of 12 isolates cluster closely. (c) *E. coli* ST1193: two pairs of samples had low SNP distances between them, but only one of these pairs suggested transmission between patients. No heatmap was produced for *E. coli* ST127 as there was no SNP difference between all five isolates.

**Table S1: ST131, ST131-like and ST5640 MLST allele profile comparison**

| Isolate | Species | MLST | Allele 1 | Allele 2 | Allele 3 | Allele 4 | Allele 5 | Allele 6 | Allele 7 |
|---|---|---|---|---|---|---|---|---|---|
| LG-4 | ECOLI | ST131 | adk(53) | fumC(40) | gyrB(47) | icd(13) | mdh(36) | purA(28) | recA(29) |
| XB-1 | ECOLI | ST- | adk(53) | fumC(40) | gyrB(47) | icd(13) | mdh(˜36) | purA(28) | recA(29) |
| MX-1 | ECOLI | ST- | adk(53) | fumC(40) | gyrB(47) | icd(13) | mdh(˜36) | purA(28) | recA(29) |
| IC-1 | ECOLI | ST5640 | adk(492) | fumC(40) | gyrB(47) | icd(13) | mdh(36) | purA(28) | recA(29) |

Two isolates were not classified by MLST, one from patient XB and another from patient MX. These two samples shared 6/7 MLST alleles with ST131, with the only differing allele predicted to be a novel full length as the corresponding allele of ST131. These two samples were manually classified as ST131-like. Moreover, one isolate from patient IC was classified as ST5640 which is also only one MLST allele different from ST131; upon further investigation this variant of the MLST allele was found to be only one SNP different from the corresponding allele of ST131. Thus, for the phylogenetic analysis, these three samples were included with the ST131 group samples.

**Table S2: Single-linkage clustering results across ST groups**

| ST group | Cluster | No. patients | Patients (no. samples) | No. samples |
|---|---|---|---|---|
| ST131 | 1 | 5 | XB (5), LG (1), MX(1), QK (1), IC(1) | 9 |
| ST131 | 2 | 1 | VN (1) | 1 |
| ST131 | 3 | 1 | YE (1) | 1 |
| ST131 | 4 | 1 | YE (1) | 1 |
| ST13 | 1 | 10 | FM (7), LG (5), QW (4), EB (3), XG (3), AZ (2), LI (2), WX (2), MY (1), XB (1) | 30 |
| ST127 | 1 | 5 | BK (1), LW (1), VB (1), VD (1), WA (1) | 5 |
| ST1193 | 1 | 1 | IA (2) | 2 |
| ST1193 | 2 | 2 | TO (1), VN (1) | 2 |

Cluster numbering is within each ST group. Overall, eight clusters were identified, four of which (one from each MLST group) contained multiple patients.
